# EXTENSIVE GENETIC INTERACTIONS (EPISTASIS) LINKED TO ALCOHOL USE DISORDER IN A HIGH-RISK POPULATION

**DOI:** 10.1101/2024.12.29.24319759

**Authors:** Stanislav Listopad, Qian Peng

## Abstract

Alcohol use disorder (AUD) is known to have a significant genetic component, yet there remains a substantial gap between its heritability and findings from genome-wide association studies. One potential factor contributing to this gap may be genetic interactions, or epistasis, a largely unexplored aspect in the context of AUD. The aim of this study was to investigate the role of epistasis in AUD susceptibility and severity among American Indians, a population that exhibits the highest rates of AUD among all ethnic groups in the U.S. We began by identifying genes previously linked to alcohol dependence and AUD, then expanded this gene set through biological networks, ultimately comprising 3,736 genes and regulatory elements. The final gene set was mapped to over 476K variants in an American Indian cohort of 742 individuals. We performed a pairwise genetic interaction association analysis on the variant set, followed by a bi-clustering procedure to group the interacting SNP pairs into interacting intervals. A total of 114 interacting pairs of genes and regulatory elements were identified to be significantly associated with AUD severity. These genes were enriched for immune system, cell adhesion, neuronal, and disease pathways. Their expressions were particularly enriched in midbrain GABAergic neurons. Our study represents the first large-scale genetic interaction study of AUD in any population. Our findings suggest that epistasis may significantly contribute to the development and progression of AUD.

## INTRODUCTION

Alcohol use disorder (AUD) is a medical condition characterized by an impaired ability to stop or control alcohol use despite experiencing adverse social, occupational, or health consequences (National Institute on Alcohol Abuse and Alcoholism, 2020). According to the 2023 National Survey on Drug Use and Health, approximately 28.9 million people aged 12 and older - representing 8.7% of the U.S. population - had AUD in the past year. Rates of AUD also vary across different populations and ethnic groups, with American Indians (AI) showing the highest prevalence in the U.S. (Substance Abuse and Mental Health Services Administration, 2023). Like many other complex diseases, the risk of developing AUD is influenced by both environmental and genetic factors, with twin studies suggesting that heritability is about 50% (Verhulst, Neale, and Kendler 2015).

With increasingly large sample sizes, meta-analyses of genome-wide association studies (GWAS) have begun to yield more robust results for AUD (Zhou et al. 2023). While GWAS is a powerful tool for genetic mapping, most of the variants identified thus far have shown only small effects, explaining a limited portion (<10%) of the AUD heritability (Zhou et al. 2023). A number of factors have been proposed to account for this issue of “missing heritability”, with genetic interactions being one potential key contributor (Eichler et al. 2010; Zuk et al. 2012; Mackay 2014). GWAS typically models genetic effects by combining multiple variants in a linear fashion, assuming additivity. However, this approach does not account for potential combinatorial effects that go beyond the linear model. When the phenotypic effects from two genetic variants deviate from the expected additive value of the individual mutations, the variants are said to exhibit a statistical epistatic interaction (Fisher 1919). Meanwhile, when two or more biological components physically interact, this is referred to as biological epistasis (Cordell 2002; Ebbert, Ridge, and Kauwe 2015). A classic example of epistasis is albinism, in which dysfunctional mutations in the gene responsible for pigment production render the pigment genes themselves irrelevant (White and Rabago-Smith 2011). Once treated as an exception, epistasis is now increasingly recognized as a common and important phenomenon in genetics (Kauffman 1993; Moore 2003). Epistasis has been observed in a variety of complex human traits and diseases, including body mass index (BMI) and Alzheimer’s disease (Mackay and Anholt 2024; Lundberg et al. 2023; D’Silva, Chakraborty, and Kahali 2022). Studies have shown that disease-related genes have a high propensity to interact with each other (Barabási, Gulbahce, and Loscalzo 2011). Furthermore, many genetic variants linked to complex diseases are located in non-coding regions, suggesting that they may exert their effects through regulatory interactions (Pickrell 2014).

Genetic interactions, however, have rarely been explored in studies on the genetics of alcohol use. Previous research into epistasis in AUD cohorts has mainly focused on a few candidate genes, such as those related to alcohol metabolic enzymes (ADH and ALDH) and dopamine receptor (DRD2) (Fernández et al. 2000; Sloan, Sayarath, and Moore 2008). The only interaction identified in relation to AUD was between *ADH1B* and *ADH7* in a Han Chinese population (Osier et al. 2004). This limited focus likely stems from the assumption that statistical power is insufficient for detecting genetic interactions on a large scale. However, it has been shown that while the number of potentially interacting gene pairs is enormous, the overall genetic interaction density is low, and not all statistical interactions are necessarily biologically meaningful (Schuldiner et al. 2005). In this light, we hypothesize that AUD-related genes are more likely to be involved in genetic interactions than previously thought. By focusing further on functionally connected genes and regulatory elements known *a priori*, we can significantly increase the statistical power to detect these interactions. We applied these insights to an American Indian cohort, which exhibits a notably high lifetime prevalence of AUD, particularly in its severe form. In doing so, we conducted the first large-scale epistasis analysis on an AUD phenotype. We began with a set of genes broadly associated with alcohol dependence and use disorders and expanded this list by leveraging biological networks. This approach led us to a comprehensive set of approximately 2,300 genes and 1,400 regulatory elements, encompassing nearly half a million SNPs. In this study, we focused on pairwise (2nd order) interactions and performed an exhaustive search for epistasis among all selected SNPs. We then applied a bi-clustering algorithm to exploit the correlations among neighboring SNPs, successfully identifying many pairs of significantly interacting genes associated with the severity of AUD.

## METHODS

### Participants

American Indian participants were recruited from eight geographically contiguous reservations. The reservations had a total population of about 3000 individuals. To be included in the study, participants had to be between the ages of 18 and 70 years, and mobile enough to be transported from their home to The Scripps Research Institute (TSRI). The demographics of the population are listed in Supplemental Table 1. Notably, the participant cohort exhibited high degree of relatedness. The protocol for the study was approved by the Institutional Review Board (IRB) of TSRI, and the board of the Indian Health Council, a tribal review group overseeing health issues for the reservations where the recruitment was undertaken. Written informed consent was obtained from each participant after the study was fully explained.

### Phenotype and genotypes

Participants were interviewed based on the Semi-Structured Assessment for the Genetics of Alcoholism (SSAGA) (Bucholz et al., 1994; Hesselbrock et al., 1999). Diagnoses of lifetime DSM-5 AUD were generated using SSAGA. The interview also retrospectively asks about the occurrence of alcohol-related life events, and the age at which the problem first occurred, from which a quantitative phenotype, the severity level of AUD, was computed. The severity level of AUD is indexed by the 36 alcohol-related life events (Supplemental Table 3) in the clinical course of the disorder, with life events given a severity weight of 1 for events 1-12; 2 for 13-24; and 3 for 25-36 (Ehlers et al. 2004, 2; Schuckit et al. 1993). AUD severity was then calculated as the sum of the severity weights of the 36 life events (Peng et al. 2019). The AUD severity phenotype is highly correlated with AUD diagnoses (Spearman’s rank correlation coefficient is 0.9, Supplemental Figure 1) (Peng et al. 2024). Additional details regarding participant recruitment and SSAGA are given in Supplemental Materials.

The whole genome sequencing pipeline of the AI cohort was previously described (Bizon et al. 2014). In brief: blood derived DNA was sequenced using Illumina low-coverage whole genome sequencing. The pair-end sequencing was performed on HiSeq2000 sequencers (Illumina). About 80% of the samples had coverage between 3X and 12X. The qualities of the variant calling were confirmed using an Affymetrix Exome1A chip. Reads from whole genome sequencing were aligned to the GRCh37/hg19 human reference genome using BMA: and realigned near indels with GATK (DePristo et al. 2011). Variants were called using both GATK Unified Genotyper following the best practices for low-coverage samples and the LD-aware variant caller Thunder (Van der Auwera et al. 2013; Li et al. 2011). Several quality control procedures were applied which resulted in a total of 10,648,122 SNPs. More details regarding sample processing and quality control can be found in Supplemental Materials.

### Core gene set

We began by identifying a set of genes that were either associated with AUD, alcohol dependence (AD), or were direct molecular targets of ethanol, which resulted in 506 genes (Supplemental Table 2). We refer to this set of genes as the *core gene set*. The sources of the core gene set are: 1) AUD associated genes in Diseases, Disgenet, and MalaCards databases (Grissa et al. 2022; Piñero et al. 2020; Rappaport et al. 2013); 2) AD associated genes in Disgenet, KEGG Disease, and MalaCards databases (Kanehisa et al. 2019); 3) genes identified by the largest GWAS meta-analysis of problematic alcohol use (Kranzler et al. 2019; Zhou et al. 2023); 4) AUD genes previously identified in various analyses of the AI cohort (Peng et al. 2017; 2019; Peng and Ehlers 2021); and 5) molecular targets of ethanol (Harris, Trudell, and Mihic 2008; Abrahao, Salinas, and Lovinger 2017). The core gene set is described in further detail within the Supplemental Materials.

### Core gene set expansion

STRING (StringDB) protein-protein interaction database and GeneHancer regulatory interaction database (the GRCh37/hg19 version) were used to expand the core gene set (Szklarczyk et al. 2023; Fishilevich et al. 2017). We refer to the resulting gene set as the *expanded core set*. StringDB was used to expand the core gene set in the following manner: for every gene in the core gene set, all of its neighbors that possessed overall score above 900, database score above 0.01, and experimental score above 0.01 were added to the expanded core set. These scores indicate high level of confidence that interaction exists and is supported by experimental and curated database evidence. For GeneHancer, any regulatory element that regulates a core gene, with a confidence score of more than 25, was added to the expanded core set. The final expanded core set contained 2285 genes, and 1451 regulatory elements. For the regulatory elements, the chromosomal loci were taken directly from GeneHancer. For genes, UCSC-hg19 genome assembly chromosomal loci were used. Two kilo byte (kb) windows were added around each gene, except for the major histocompatibility complex genes that have multiple alternative loci. The UCSC-hg19 genome assembly was obtained from UCSC genome browser table browser (Supplementary Files 1 and 2) (Nassar et al. 2023). We then selected all the SNPs within the AI dataset that belonged to genes and/or regulatory elements within the expanded core set and had minor allele frequency higher than 0.01, resulting in 476,850 SNPs.

### Epistasis analysis

The SNP x SNP pairwise interaction analysis was performed using rapid epistatic mixed model association analysis (REMMA). REMMA is a linear mixed model tool to exhaustively search for pairwise interactions while accounting for both population stratification and individual relatedness (Ning et al. 2018; Wang et al. 2020). An exhaustive algorithm, albeit more computationally intensive, was chosen due to concerns about the performance of non-exhaustive algorithms (Ponte-Fernández et al. 2022). We performed the epistasis analysis using five different REMMA configurations and then merged the results (Supplemental Table 7). Sex and age were included as covariates. Further details are given in the Supplementary Materials.

### Bi-clustering of SNP x SNP epistasis analysis results

Due to linkage disequilibrium (LD), the effects of SNPs located near each other are often correlated. Therefore, we employed a bi-clustering algorithm to group the SNP pairs obtained from REMMA (Hannum et al. 2009). The algorithm clusters interacting SNP pairs into contiguous interacting SNP *intervals* that are enriched for interacting SNP pairs. For the bi-clustering algorithm, every SNP pair was considered as either interacting or non-interacting by thresholding on its REMMA p-value. We chose p-value of < 1e-8 to transform the REMMA results into a set of interacting SNP pairs. Note that selection of the p-value threshold impacts the bi-clustering results and their interpretations. We performed bi-clustering on each chromosome pair individually to distribute the analysis. To illustrate the process, interactions can be represented as a matrix. SNPs from one chromosome are rows, while SNPs from the other chromosome are columns of an interaction matrix (Figure 1). The values in the matrix are either ones (interacting SNP pairs) or zeros (no interaction). A sampling matrix of varying size, from 1x1 to the maximum of 60x60 is then traversed across the entirety of the interaction matrix. The maximum sampling matrix size contributes significantly toward algorithm runtime. Each instance of the sampling matrix is evaluated for interaction enrichment using hypergeometric distribution. P-values from the hypergeometric tests are reported. The maximum number of the hypergeometric tests performed for the entire analysis is bounded by the number of rows (A), in the interaction matrix, times the number of columns (B) times the maximum size of the sampling matrix. To compute the bi-clustering p-value threshold using Bonferroni correction, assume that A and B both equal the total number of SNPs. The significance threshold is then set to 0.05 / (A*B*60^2^). A less conservative threshold can be obtained by tracking the number of hypergeometric tests performed within the bi-clustering procedure. Note that while the resulting clusters are enriched for interacting SNPs, they also contain some non-interacting SNPs. The interacting SNPs within each significant interacting interval were then mapped to genes and/or regulatory elements that they were located within. Thus, pairs of interacting intervals were translated into interacting gene and/or regulatory element pairs.

**Figure 1:**
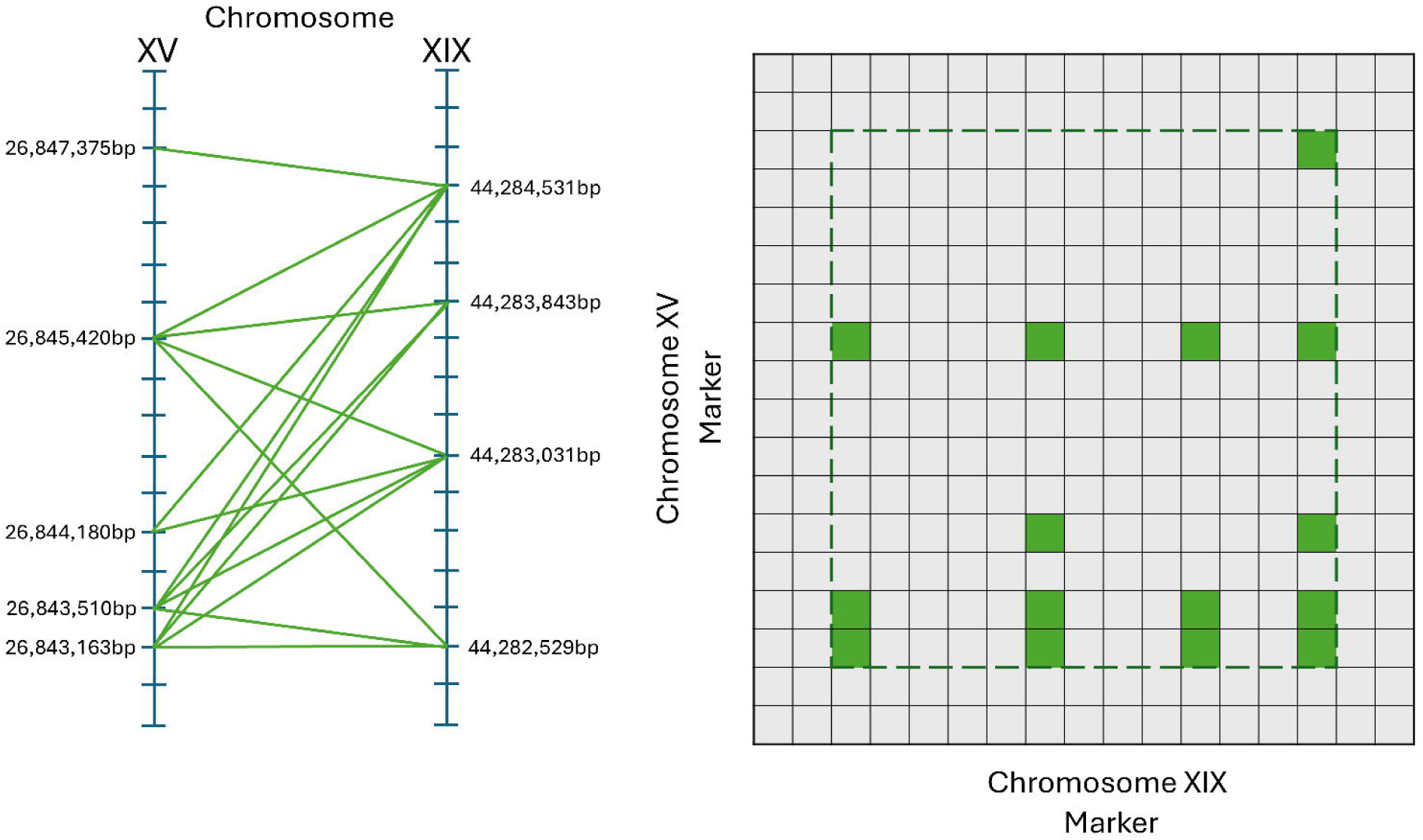
Interacting SNP pairs are grouped into interacting SNP-intervals using a bi-clustering algorithm. In this illustration, 15 pairs of nearby SNP interactions between chromosomes 15 and 19 are clustered into one 14x13 interval pair. The interacting SNPs on chromosome 15 belong to gene GABRB3, and the interacting SNPs on chromosome 19 belong to gene KCNN4.

### Analysis of interacting genes and regulatory elements

The bi-clustering results contained two types of significantly interacting pairs: gene x gene and gene x regulatory element. For each such pair, we looked for previous evidence of possible interaction using: StringDB, GeneHancer, MSigDB curated gene sets, and literature search. Details can be found in Supplemental Materials. We also conducted two pathway and functional enrichment analyses on the significantly interacting pairs. We performed enrichment analyses of all the interacting genes using the expanded core gene set as the background. Additionally, we assembled all the protein coding genes regulated by the interacting regulatory elements with GeneHancer confidence score greater than 25 and then conducted enrichment analysis of these genes using the entire human gene set as the background.

## RESULTS

### Interacting SNP pairs

Out of 476,850 SNPs analyzed, 150,676 could be considered effectively independent due to LD structure (Gao 2011). Using the number of effectively independent SNPs, we set p-value at 4.4e-12 as the significance threshold for all SNP x SNP interactions using Bonferroni correction. Note that the correction is overly conservative, as the tested pairs are not all independent of each other, even though linkage disequilibrium has already been accounted for. None of the interacting SNP pairs passed this significance threshold. Several SNP pairs were suggestively significant at p-value < 4.4e-11 (Table 1). The top interacting SNPs are between one SNP on the insulin receptor gene (*INSR*) and multiple SNPs located on the cholinergic receptor nicotinic gamma subunit gene (*CHRNG*). For the bi-clustering stage, we used p-value < 1e-8 as threshold, which resulted in a total of 1539 interacting SNP pairs. These interacting SNP pairs were spread out across 84 chromosome pairs, such as chromosome 1 – chromosome 1 pair, chromosome 1 – chromosome 2 pair, and etc.

**Table 1:**
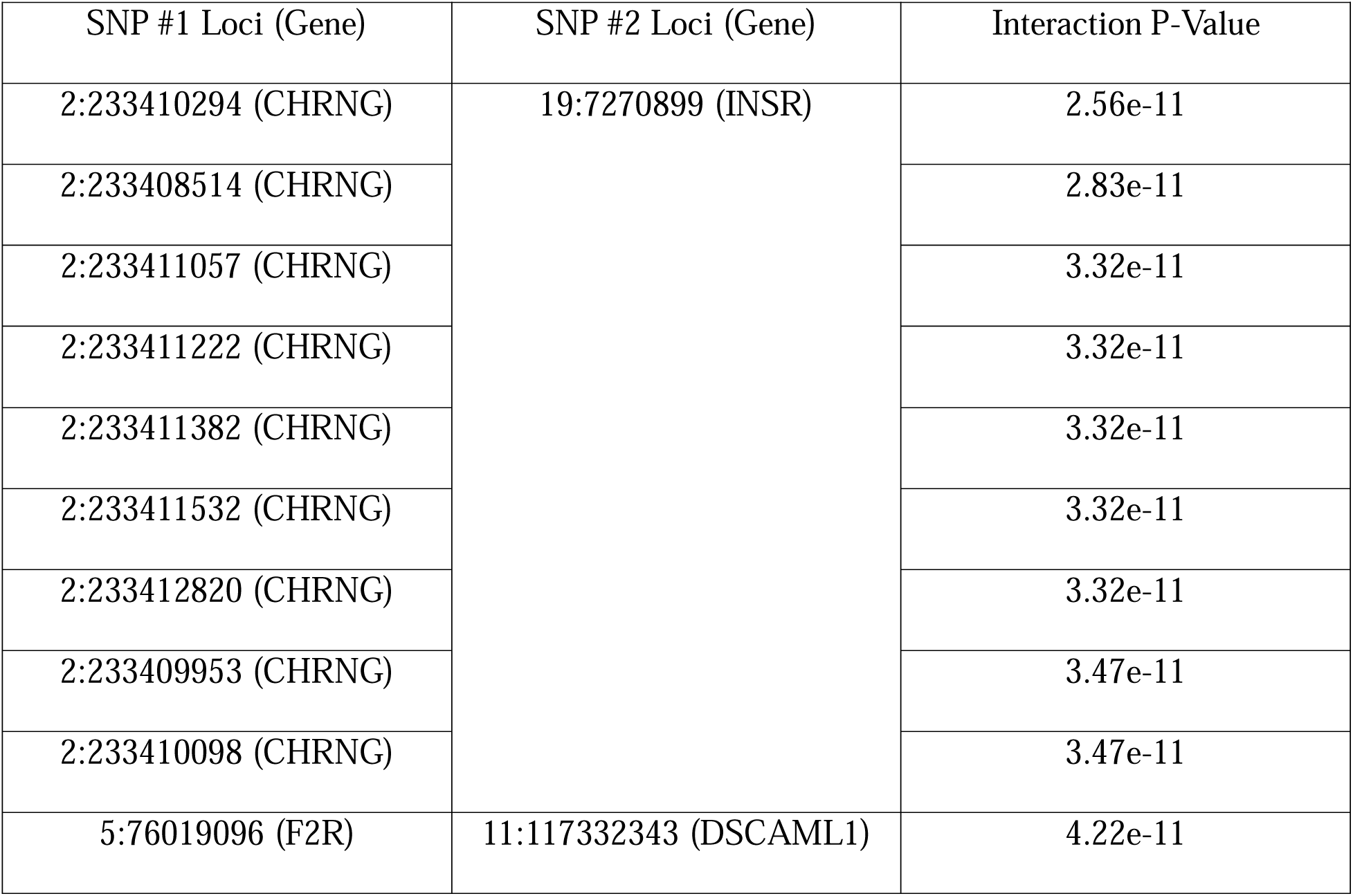
Top (p-value < 4.4e-11) interacting SNP pairs according to REMMA epistasis analysis.

### Significantly interacting gene and regulatory element pairs

We have identified a total of 114 interacting pairs of genes and/or regulatory elements to be significantly associated with AUD severity after bi-clustering REMMA results (Figure 2, Supplemental Table 4). Of these, 7 pairs contained at least one regulatory element, while 107 were gene pairs. Of 160 genes and 6 regulatory elements involved in the significant interactions, 58 have been previously connected to alcohol use disorder. About half of the interacting pairs form small networks together with other interacting pairs. The largest interacting network contains 12 genes. For 30 gene and/or regulatory element pairs there existed prior evidence of potential biological interactions in scientific literature and/or pathway databases. Several interacting pairs were present within same neuronal system pathways such as Reactome’s transmission across chemical synapses (Milacic et al. 2024). The rest were present in various disease, immune system, and signaling pathways. The interacting pairs were distributed across all chromosomes except chromosome 22 (Figure 3).

**Figure 2:**
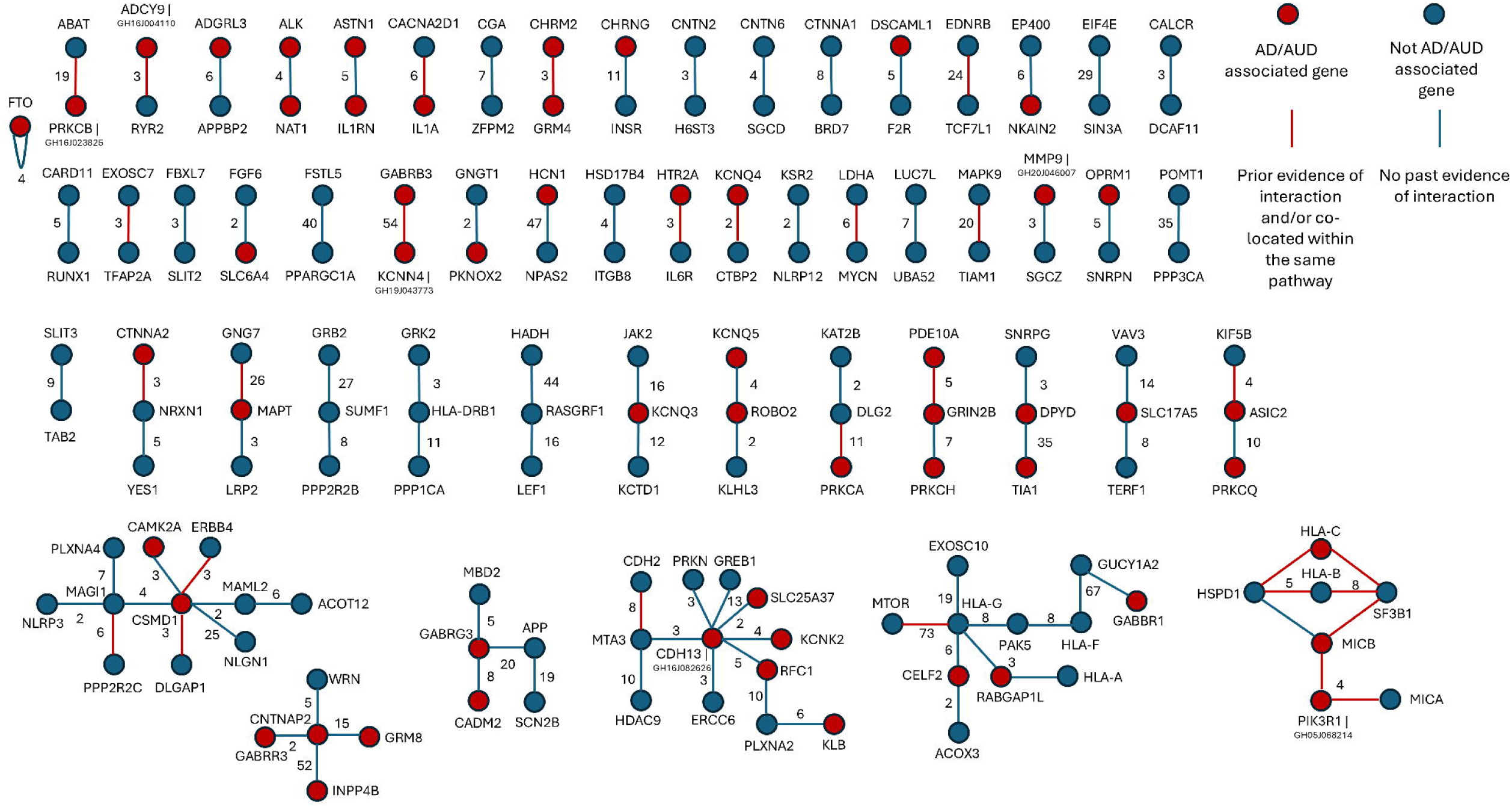
Interacting pairs of genes and/or regulatory elements that were significantly associated with AUD severity in the American Indian cohort. Interactions between pairs of genes and/or regulatory elements are represented as line edges between circle nodes. Each node is labeled with the name of the gene and/or regulatory element. Red nodes (part of the core gene set) have been associated with alcohol dependence or alcohol use disorder in the past, there is no such evidence for blue nodes (part of the expanded core gene set). Red edges represent interactions for which there has been some evidence in the past, such as being located within the same pathway; no previous evidence was found for blue edges. The numbers next to the edges represent the total number of interacting SNP pairs belonging to the given pair of genes and/or regulatory elements. A small number of nodes represents multiple overlapping genes, all part of major histocompatibility complex (MHC) on chromosome six. In such cases the overlapping genes are listed as follows: gene1|gene2|gene3|etc. Additionally, all regulatory elements share intervals with genes.

**Figure 3:**
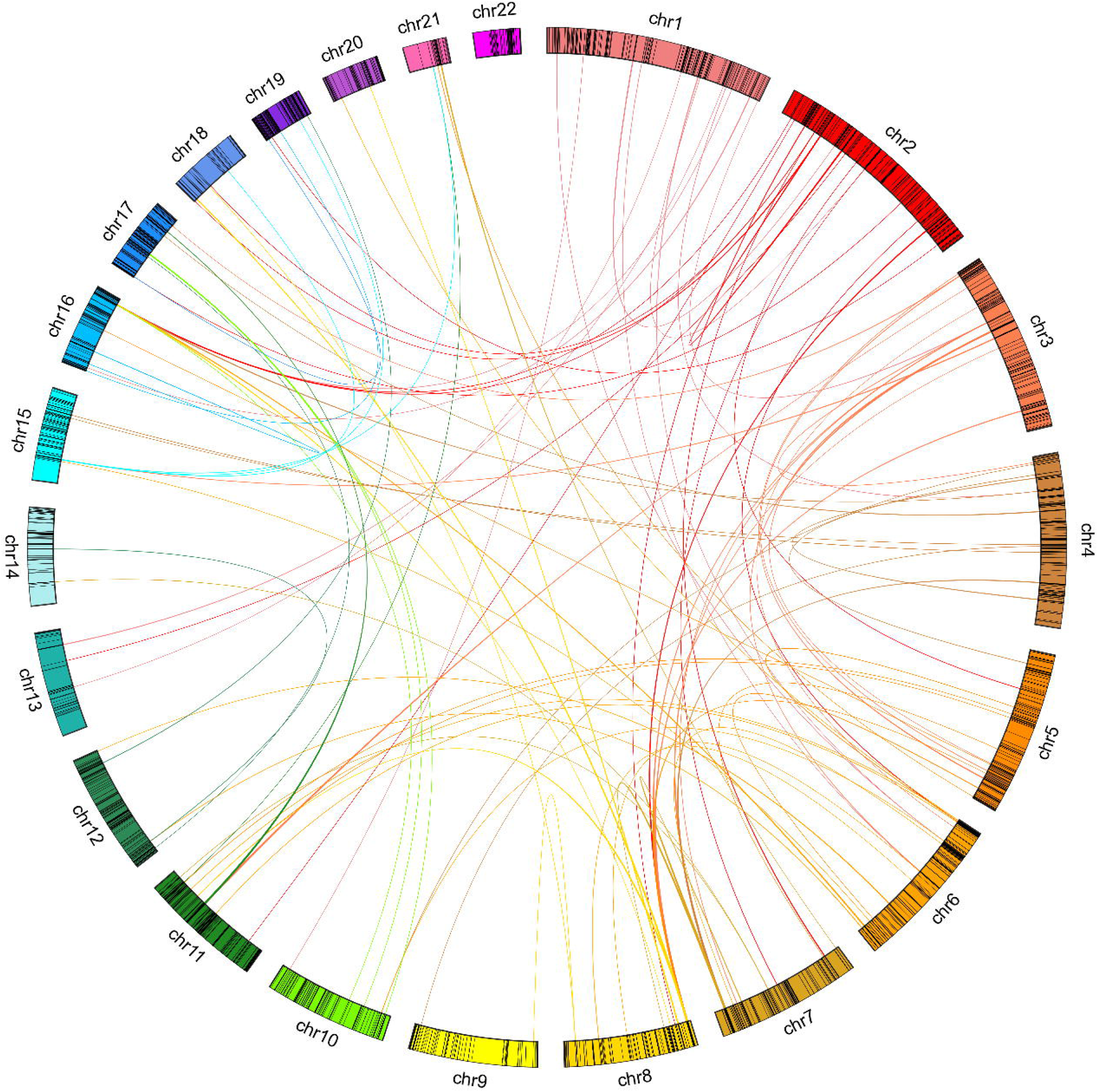
A Circos plot of interacting gene and/or regulatory element pairs significantly associated with AUD severity in the Ameican Indian cohort across all human chromosomes. The black lines on chromosomes correspond to the expanded core gene set.

### Pathway enrichment analysis of interacting genes

The significantly interacting gene pairs were enriched for, cell adhesion molecule (CAM), disease, immune system, and neuronal pathways (Supplemental Table 5). CAMs play a role in a variety of processes including homeostasis, immune response, inflammation, and development of neuronal tissue. CAMs have also been implicated in addiction due to their impact on neuroplasticity (Muskiewicz, Uhl, and Hall 2018; Zhong et al. 2015; Levchuk et al. 2020). The enriched disease pathways encompassed autoimmune thyroid disease, diabetes, and graft rejection pathways. The immune system pathways belonged to three categories: cytotoxicity regulation, major histocompatibility complex, and T cell function. Immune system is known to play a role in AUD due to its impact on neuroinflammation (Crews 2012; Cui, Shurtleff, and Harris 2014; Lucerne and Kiraly 2021). Additionally, the tissue specificity analysis demonstrated significant upregulation of genes for most brain regions including frontal cortex, anterior cingulate cortex, basal ganglia, hypothalamus, hippocampus and amygdala (Supplemental Table 5, Supplemental Figure 2). The cell type analysis identified enrichment for midbrain GABAergic neurons and neuroblasts and fetal lung visceral neurons (Supplemental Figure 3). The genes regulated by interacting regulatory elements were enriched for gene transcription, cancer, neuronal, and various signaling pathways (Supplemental Table 6).

## DISCUSSION

Understanding gene interactions is crucial to deciphering the structure and function of complex genetic systems that contribute to diseases. The first step in this process is identifying the scope of these interactions. To achieve this, we performed a large-scale epistasis analysis to investigate the genetic factors associated with AUD severity in an American Indian cohort, which has a notably high AUD prevalence of 70%, with half of those affected experiencing severe AUD. This is the first study of its kind examining alcohol use or use disorder in any population.

### Extensive genetic interactions linked to AUD severity

We identified a total of 114 interactions involving 160 genes and 6 regulatory elements that are significantly associated with the AUD severity phenotype in this AI population. These interactions may occur across various biological contexts. While statistical epistasis does not always directly translate into biological epistasis, many of the epistatic pairs we identified have been previously proposed to interact biologically, either directly or through a few intermediaries. Notably, five of the gene pairs we found to interact had been previously suggested in the literature as potentially interacting: *ASIC2* x *KIF5B*, *CSMD1* x *ERBB4*, *MTA3* x *CDH2*, *MYCN* x *LDHA*, and *SF3B1* x *HLA-B* (Gialeli et al. 2021; Xu et al. 2016; Ahmed et al. 2021; Dorneburg et al. 2018; Biernacki et al. 2020). The *ASIC2* and *KIF5B* may interact indirectly via Syntabulin. Meanwhile, *CSMD1* may regulate epidermal growth factors, of which *ERBB4* is a member. Another two gene pairs, *CSMD1* x *DLGAP1* and *NRXN1* x *CTNNA2*, have been implicated as interacting by the protein-protein interaction database StringDB. We did not replicate the *ADH1B* x *ADH7* interaction previously identified in relation to AUD in the Han Chinese population (Osier et al. 2004). This is not surprising, as the *ADH1B* allele rs1229984 implicated in the reported interaction has a MAF of 26% in the Han Chinese population (Chang et al. 2023) but only 3% in our American Indian cohort.

Over a quarter of the interacting gene pairs are co-located within various biological pathways, suggesting potential mechanisms of interaction. The most common pathways involve neuronal pathways, followed by signaling pathways, human disease pathways, and immune system pathways. Notably, tissue-and cell-type specific analyses revealed that the interacting genes were most significantly enriched in the frontal cortex, anterior cingulate cortex, caudate, nucleus accumbens, and developmental midbrain GABAergic neurons, which play key roles in the development and progression of alcohol and substance use disorders. Interacting gene pairs that show both statistical and biological evidence of epistasis may represent particularly promising candidates for further investigation in future studies of AUD.

### Strengths and Limitations

To overcome the limitations of statistical power in SNP-level epistasis analysis and to enhance the interpretability of the results, we adopted a strategy that integrated both hypothesis-driven and discovery-driven approaches. By prioritizing nearly half a million SNPs located in genes and regulatory elements linked to alcohol use and use disorders—both directly and indirectly—and accounting for SNP correlations, we were able to achieve significant findings, even with a moderate sample size.

The results of this study should be interpreted with several limitations in mind, however. First, the analyses were not designed to generate a comprehensive model of AUD in this community but rather to explore specific genetic interactions associated with AUD phenotypes. As such, a larger sample size, powered to assess genome-wide epistasis, would be necessary for more robust conclusions. Additionally, American Indians represent a diverse group both genetically and environmentally, meaning our findings may not be applicable to other American Indian populations. Prevalence of AUD, for instance, varies among tribes, which may limit the generalizability of our results (Beals et al. 2005; Ehlers et al. 2004). This diversity however underscores the importance of identifying risk and resilience factors within specific communities, as these insights can help guide targeted prevention and intervention efforts. Moreover, our study relied on both genotype data and extensive phenotyping instrument, but no replication sample of American Indians is currently available. Given that alcohol and substance use risks in American Indian populations are generally understudied, this gap in research highlights the need for further exploration. While our specific findings may not be generalizable to other populations, the methodology used and the gene interactions identified for AUD in this study may be highly relevant to other groups, warranting further investigations.

In summary, this study represents the first large-scale investigation into epistasis in the context of alcohol use disorders. By systematically studying how genetic interactions influence AUD traits and expanding the boundaries of traditional GWAS, we open the door to new insights into how genes work together and interact to drive the disorder, offering a more nuanced understanding of the complex genetic architecture of AUD that could lead to more effective prevention and treatment strategies. While some of our findings may be specific to an American Indian population, overall, our results suggest that genetic interactions could play a significant role in the development and progression of AUD.

## Supporting information

Supplemental Results and Methods

## Data Availability

The data that support the finding of this study are available by contacting the last author. However, data availability is subject to approval of the specific American Indian tribes participating. Code used to perform core gene set expansion, epistasis analysis, bi-clustering, and other miscellaneous tasks can be found in our GitHub repository [https://github.com/staslist/Biclustering_Epistasis]. The code in the repository contains ready to use bi-clustering functionality. REMMA (version 2021.6.17) can be downloaded from Python pip package installer.

## Ethical Approval

The authors assert that all procedures and protocols for the study were carried out in accordance with the latest version of the Declaration of Helsinki. The protocol and procedures were approved by the Institutional Review Board of The Scripps Research Institute and Indian Health Council for the AI tribes participating. Written informed consent was obtained from each participant after the study was fully explained by study staff.

## ACKNOWLEDGEMENTS

We would like to acknowledge and thank all of our American Indian participants, and the following people for their roles in 1) the genotyping effort: Kirk Wilhelmsen, Scott Chasse, Piotr Mieczkowski, Ewa Patrycja Malc, Joshua Sailsbery, Phil Owens, and Chris Bizon; 2) recruiting participants, and collection and preparation of the clinical data: Cindy Ehlers, David Gilder, Corinne Kim, Evie Phillips, Phillip Lau, and Derek Wills; and 3) discussion on ethanol molecular targets: Candice Contet.

This work was supported by the National Institutes of Health (NIH): National Institute on Alcohol Abuse and Alcoholism (NIAAA) T32AA007456 to SL, and National Institute on Drug Abuse (NIDA) DP1DA054373 to QP. NIAAA and NIDA had no further role in the study design; in the collection, analysis and interpretation of data; in the writing of the report; or in the decision to submit the article for publication. The authors report no conflicts of interest, competing interests or undisclosed financial support. The authors alone are responsible for the content and writing of this paper, and the views expressed do not necessarily represent the views of the NIH, NIAAA or NIDA.

## Citations

Abrahao, Karina P., Armando G. Salinas, and David M. Lovinger. 2017. “Alcohol and the Brain: Neuronal Molecular Targets, Synapses, and Circuits.” Neuron 96 (6): 1223–38. 10.1016/j.neuron.2017.10.032.

Ahmed, Mahmoud, Trang Huyen Lai, Wanil Kim, and Deok Ryong Kim. 2021. “A Functional Network Model of the Metastasis Suppressor PEBP1/RKIP and Its Regulators in Breast Cancer Cells.” Cancers 13 (23): 6098. 10.3390/cancers13236098.

Barabási, Albert-László, Natali Gulbahce, and Joseph Loscalzo. 2011. “Network Medicine: A Network-Based Approach to Human Disease.” Nature Reviews Genetics 12 (1): 56–68. 10.1038/nrg2918.

Beals, Janette, Douglas K. Novins, Nancy R. Whitesell, Paul Spicer, Christina M. Mitchell, Spero M. Manson, and American Indian Service Utilization, Psychiatric Epidemiology, Risk and Protective Factors Project Team. 2005. “Prevalence of Mental Disorders and Utilization of Mental Health Services in Two American Indian Reservation Populations: Mental Health Disparities in a National Context.” American Journal of Psychiatry 162 (9): 1723–32. 10.1176/appi.ajp.162.9.1723.

Biernacki, Melinda Ann, Kimberly A Foster, Courtnee Clough, Stephanie Busch, Carrie Cummings, Vivian G. Oehler, Derek Stirewalt, Sergei Doulatov, and Marie Bleakley. 2020. “A Shared SF3B1 Neoantigen Is Presented on Primary Malignant Cells and Induced Pluripotent Stem Cell-Derived Hematopoietic Lines.” Blood 136 (November):13–14. 10.1182/blood-2020-139907.

Bizon, Chris, Michael Spiegel, Scott A. Chasse, Ian R. Gizer, Yun Li, Ewa P. Malc, Piotr A. Mieczkowski, et al. 2014. “Variant Calling in Low-Coverage Whole Genome Sequencing of a Native American Population Sample.” BMC Genomics 15 (1): 85. 10.1186/1471-2164-15-85.

Chang, Ting-Gang, Ting-Ting Yen, Chia-Yi Wei, Tzu-Hung Hsiao, and I-Chieh Chen. 2023. “Impacts of Rs1229984 and Rs671 Polymorphisms on Risks of Alcohol-Related Disorder and Cancer.” Cancer Medicine 12 (1): 747–59. 10.1002/cam4.4920.

Cordell, Heather J. 2002. “Epistasis: What It Means, What It Doesn’t Mean, and Statistical Methods to Detect It in Humans.” Human Molecular Genetics 11 (20): 2463–68. 10.1093/hmg/11.20.2463.

Crews, Fulton T. 2012. “Immune Function Genes, Genetics, and the Neurobiology of Addiction.” Alcohol Researchl: Current Reviews 34 (3): 355–61.

Cui, Changhai, David Shurtleff, and R. Adron Harris. 2014. “Chapter One - Neuroimmune Mechanisms of Alcohol and Drug Addiction.” In International Review of Neurobiology, edited by Changhai Cui, David Shurtleff, and R. Adron Harris, 118:1–12. Neuroimmune Signaling in Drug Actions and Addictions. Academic Press. 10.1016/B978-0-12-801284-0.00001-4.

DePristo, Mark A., Eric Banks, Ryan Poplin, Kiran V. Garimella, Jared R. Maguire, Christopher Hartl, Anthony A. Philippakis, et al. 2011. “A Framework for Variation Discovery and Genotyping Using Next-Generation DNA Sequencing Data.” Nature Genetics 43 (5): 491–98. 10.1038/ng.806.

Dorneburg, Carmen, Matthias Fischer, Thomas F.E. Barth, Wolfgang Mueller-Klieser, Barbara Hero, Judith Gecht, Daniel R. Carter, et al. 2018. “LDHA in Neuroblastoma Is Associated with Poor Outcome and Its Depletion Decreases Neuroblastoma Growth Independent of Aerobic Glycolysis.” Clinical Cancer Research 24 (22): 5772–83. 10.1158/1078-0432.CCR-17-2578.

D’Silva, Sheldon, Shreya Chakraborty, and Bratati Kahali. 2022. “Concurrent Outcomes from Multiple Approaches of Epistasis Analysis for Human Body Mass Index Associated Loci Provide Insights into Obesity Biology.” Scientific Reports 12 (1): 7306. 10.1038/s41598-022-11270-0.

Ebbert, Mark T. W., Perry G. Ridge, and John S. K. Kauwe. 2015. “Bridging the Gap between Statistical and Biological Epistasis in Alzheimer’s Disease.” BioMed Research International 2015 (1): 870123. 10.1155/2015/870123.

Ehlers, Cindy L., Tamara L. Wall, Michelle Betancourt, and David A. Gilder. 2004. “The Clinical Course of Alcoholism in 243 Mission Indians.” American Journal of Psychiatry 161 (7): 1204–10. 10.1176/appi.ajp.161.7.1204.

Eichler, Evan E., Jonathan Flint, Greg Gibson, Augustine Kong, Suzanne M. Leal, Jason H. Moore, and Joseph H. Nadeau. 2010. “Missing Heritability and Strategies for Finding the Underlying Causes of Complex Disease.” Nature Reviews Genetics 11 (6): 446–50. 10.1038/nrg2809.

Fernández, José R., Lisa M. Tarantino, Scott M. Hofer, George P. Vogler, and Gerald E. McClearn. 2000. “Epistatic Quantitative Trait Loci for Alcohol Preference in Mice.” Behavior Genetics 30 (6): 431–37. 10.1023/A:1010232900342.

Fisher, R. A. 1919. “The Causes of Human Variability.” The Eugenics Review 10 (4): 213–20.

Fishilevich, Simon, Ron Nudel, Noa Rappaport, Rotem Hadar, Inbar Plaschkes, Tsippi Iny Stein, Naomi Rosen, et al. 2017. “GeneHancer: Genome-Wide Integration of Enhancers and Target Genes in GeneCards.” Database 2017 (January):bax028. 10.1093/database/bax028.

Gao, Xiaoyi. 2011. “Multiple Testing Corrections for Imputed SNPs.” Genetic Epidemiology 35 (3): 154–58. 10.1002/gepi.20563.

Gialeli, Chrysostomi, Emre Can Tuysuz, Johan Staaf, Safia Guleed, Veronika Paciorek, Matthias Mörgelin, Konstantinos S. Papadakos, and Anna M. Blom. 2021. “Complement Inhibitor CSMD1 Modulates Epidermal Growth Factor Receptor Oncogenic Signaling and Sensitizes Breast Cancer Cells to Chemotherapy.” Journal of Experimental & Clinical Cancer Researchl: CR 40 (August):258. 10.1186/s13046-021-02042-1.

Grissa, Dhouha, Alexander Junge, Tudor I Oprea, and Lars Juhl Jensen. 2022. “Diseases 2.0: A Weekly Updated Database of Disease–Gene Associations from Text Mining and Data Integration.” Database 2022 (January):baac019. 10.1093/database/baac019.

Hannum, Gregory, Rohith Srivas, Aude Guénolé, Haico van Attikum, Nevan J. Krogan, Richard M. Karp, and Trey Ideker. 2009. “Genome-Wide Association Data Reveal a Global Map of Genetic Interactions among Protein Complexes.” PLOS Genetics 5 (12): e1000782. 10.1371/journal.pgen.1000782.

Harris, R. Adron, James R. Trudell, and S. John Mihic. 2008. “Ethanol’s Molecular Targets.” Science Signaling 1 (28): re7–re7. 10.1126/scisignal.128re7.

Kanehisa, Minoru, Yoko Sato, Miho Furumichi, Kanae Morishima, and Mao Tanabe. 2019. “New Approach for Understanding Genome Variations in KEGG.” Nucleic Acids Research 47 (D1): D590–95. 10.1093/nar/gky962.

Kauffman, Stuart A. 1993. The Origins of Order: Self-Organization and Selection in Evolution. Oxford University Press.

Kranzler, Henry R., Hang Zhou, Rachel L. Kember, Rachel Vickers Smith, Amy C. Justice, Scott Damrauer, Philip S. Tsao, et al. 2019. “Genome-Wide Association Study of Alcohol Consumption and Use Disorder in 274,424 Individuals from Multiple Populations.” Nature Communications 10 (1): 1499. 10.1038/s41467-019-09480-8.

Levchuk, Lyudmila A., Elise M. G. Meeder, Olga V. Roschina, Anton J. M. Loonen, Anastasiia S. Boiko, Ekaterina V. Michalitskaya, Elena V. Epimakhova, et al. 2020. “Exploring Brain Derived Neurotrophic Factor and Cell Adhesion Molecules as Biomarkers for the Transdiagnostic Symptom Anhedonia in Alcohol Use Disorder and Comorbid Depression.” Frontiers in Psychiatry 11 (April). 10.3389/fpsyt.2020.00296.

Li, Yun, Carlo Sidore, Hyun Min Kang, Michael Boehnke, and Gonçalo R. Abecasis. 2011. “Low-Coverage Sequencing: Implications for Design of Complex Trait Association Studies.” Genome Research 21 (6): 940–51. 10.1101/gr.117259.110.

Lucerne, Kelsey E., and Drew D. Kiraly. 2021. “Chapter Seven -The Role of Gut-Immune-Brain Signaling in Substance Use Disorders.” In International Review of Neurobiology, edited by Erin S. Calipari and Nicholas W. Gilpin, 157:311–70. Neurobiology of Addiction and Co-Morbid Disorders. Academic Press. 10.1016/bs.irn.2020.09.005.

Lundberg, Mischa, Letitia M. F. Sng, Piotr Szul, Rob Dunne, Arash Bayat, Samantha C. Burnham, Denis C. Bauer, and Natalie A. Twine. 2023. “Novel Alzheimer’s Disease Genes and Epistasis Identified Using Machine Learning GWAS Platform.” Scientific Reports 13 (1): 17662. 10.1038/s41598-023-44378-y.

Mackay, Trudy F. C. 2014. “Epistasis and Quantitative Traits: Using Model Organisms to Study Gene–Gene Interactions.” Nature Reviews Genetics 15 (1): 22–33. 10.1038/nrg3627.

Mackay, Trudy F. C., and Robert R. H. Anholt. 2024. “Pleiotropy, Epistasis and the Genetic Architecture of Quantitative Traits.” Nature Reviews Genetics 25 (9): 639–57. 10.1038/s41576-024-00711-3.

Milacic, Marija, Deidre Beavers, Patrick Conley, Chuqiao Gong, Marc Gillespie, Johannes Griss, Robin Haw, et al. 2024. “The Reactome Pathway Knowledgebase 2024.” Nucleic Acids Research 52 (D1): D672–78. 10.1093/nar/gkad1025.

Moore, Jason H. 2003. “The Ubiquitous Nature of Epistasis in Determining Susceptibility to Common Human Diseases.” Human Heredity 56 (1–3): 73–82. 10.1159/000073735.

Muskiewicz, Dawn E., George R. Uhl, and F. Scott Hall. 2018. “The Role of Cell Adhesion Molecule Genes Regulating Neuroplasticity in Addiction.” Neural Plasticity 2018 (1): 9803764. 10.1155/2018/9803764.

Nassar, Luis R, Galt P Barber, Anna Benet-Pagès, Jonathan Casper, Hiram Clawson, Mark Diekhans, Clay Fischer, et al. 2023. “The UCSC Genome Browser Database: 2023 Update.” Nucleic Acids Research 51 (D1): D1188–95. 10.1093/nar/gkac1072.

Ning, Chao, Dan Wang, Huimin Kang, Raphael Mrode, Lei Zhou, Shizhong Xu, and Jian-Feng Liu. 2018. “A Rapid Epistatic Mixed-Model Association Analysis by Linear Retransformations of Genomic Estimated Values.” Bioinformatics 34 (11): 1817–25. 10.1093/bioinformatics/bty017.

Osier, M.v., R.-B. Lu, A.j. Pakstis, J.r. Kidd, S.-Y. Huang, and Kenneth K. Kidd. 2004. “Possible Epistatic Role of ADH7 in the Protection against Alcoholism.” American Journal of Medical Genetics Part B: Neuropsychiatric Genetics 126B (1): 19–22. 10.1002/ajmg.b.20136.

Peng, Qian, Chris Bizon, Ian R. Gizer, Kirk C. Wilhelmsen, and Cindy L. Ehlers. 2019. “Genetic Loci for Alcohol-Related Life Events and Substance-Induced Affective Symptoms: Indexing the ‘Dark Side’ of Addiction.” Translational Psychiatry 9 (1): 1–12. 10.1038/s41398-019-0397-6.

Peng, Qian, and Cindy L. Ehlers. 2021. “Long Tracks of Homozygosity Predict the Severity of Alcohol Use Disorders in an American Indian Population.” Molecular Psychiatry 26 (6): 2200– 2211. 10.1038/s41380-020-00989-9.

Peng, Qian, David A. Gilder, Rebecca A. Bernert, Katherine J. Karriker-Jaffe, and Cindy L. Ehlers. 2024. “Genetic Factors Associated with Suicidal Behaviors and Alcohol Use Disorders in an American Indian Population.” Molecular Psychiatry 29 (4): 902–13. 10.1038/s41380-023-02379-3.

Peng, Qian, Ian R. Gizer, Kirk C. Wilhelmsen, and Cindy L. Ehlers. 2017. “Associations Between Genomic Variants in Alcohol Dehydrogenase Genes and Alcohol Symptomatology in American Indians and European Americans: Distinctions and Convergence.” Alcoholism: Clinical and Experimental Research 41 (10): 1695–1704. 10.1111/acer.13480.

Pickrell, Joseph K. 2014. “Joint Analysis of Functional Genomic Data and Genome-Wide Association Studies of 18 Human Traits.” The American Journal of Human Genetics 94 (4): 559–73. 10.1016/j.ajhg.2014.03.004.

Piñero, Janet, Juan Manuel Ramírez-Anguita, Josep Saüch-Pitarch, Francesco Ronzano, Emilio Centeno, Ferran Sanz, and Laura I Furlong. 2020. “The DisGeNET Knowledge Platform for Disease Genomics: 2019 Update.” Nucleic Acids Research 48 (D1): D845–55. 10.1093/nar/gkz1021.

Ponte-Fernández, Christian, Jorge González-Domínguez, Antonio Carvajal-Rodríguez, and María J. Martín. 2022. “Evaluation of Existing Methods for High-Order Epistasis Detection.” IEEE/ACM Transactions on Computational Biology and Bioinformatics 19 (2): 912–26. 10.1109/TCBB.2020.3030312.

Rappaport, Noa, Noam Nativ, Gil Stelzer, Michal Twik, Yaron Guan-Golan, Tsippi Iny Stein, Iris Bahir, et al. 2013. “MalaCards: An Integrated Compendium for Diseases and Their Annotation.” Database 2013 (January):bat018. 10.1093/database/bat018.

Schuckit, M. A., T. L. Smith, R. Anthenelli, and M. Irwin. 1993. “Clinical Course of Alcoholism in 636 Male Inpatients.” The American Journal of Psychiatry 150 (5): 786–92. 10.1176/ajp.150.5.786.

Schuldiner, Maya, Sean R. Collins, Natalie J. Thompson, Vladimir Denic, Arunashree Bhamidipati, Thanuja Punna, Jan Ihmels, et al. 2005. “Exploration of the Function and Organization of the Yeast Early Secretory Pathway through an Epistatic Miniarray Profile.” Cell 123 (3): 507–19. 10.1016/j.cell.2005.08.031.

Sloan, Chantel D., Vicki Sayarath, and Jason H. Moore. 2008. “Systems Genetics of Alcoholism.” Alcohol Research & Health 31 (1): 14–25.

Szklarczyk, Damian, Rebecca Kirsch, Mikaela Koutrouli, Katerina Nastou, Farrokh Mehryary, Radja Hachilif, Annika L Gable, et al. 2023. “The STRING Database in 2023: Protein–Protein Association Networks and Functional Enrichment Analyses for Any Sequenced Genome of Interest.” Nucleic Acids Research 51 (D1): D638–46. 10.1093/nar/gkac1000.

Van der Auwera, Geraldine A., Mauricio O. Carneiro, Chris Hartl, Ryan Poplin, Guillermo del Angel, Ami Levy-Moonshine, Tadeusz Jordan, et al. 2013. “From FastQ Data to High Confidence Variant Calls: The Genome Analysis Toolkit Best Practices Pipeline.” *Current Protocols in Bioinformatics / Editoral Board*, Andreas D. Baxevanis … [et Al.] 11 (1110): 11.10.1–11.10.33. 10.1002/0471250953.bi1110s43.

Verhulst, B., M. C. Neale, and K. S. Kendler. 2015. “The Heritability of Alcohol Use Disorders: A Meta-Analysis of Twin and Adoption Studies.” Psychological Medicine 45 (5): 1061–72. 10.1017/S0033291714002165.

Wang, Dan, Hui Tang, Jian-Feng Liu, Shizhong Xu, Qin Zhang, and Chao Ning. 2020. “Rapid Epistatic Mixed-Model Association Studies by Controlling Multiple Polygenic Effects.” Bioinformatics 36 (19): 4833–37. 10.1093/bioinformatics/btaa610.

White, Désirée, and Montserrat Rabago-Smith. 2011. “Genotype–Phenotype Associations and Human Eye Color.” Journal of Human Genetics 56 (1): 5–7. 10.1038/jhg.2010.126.

Xu, Junyu, Na Wang, Jian-hong Luo, and Jun Xia. 2016. “Syntabulin Regulates the Trafficking of PICK1-Containing Vesicles in Neurons.” Scientific Reports 6 (1): 20924. 10.1038/srep20924.

Zhong, Xiaoming, Jana Drgonova, Chuan-Yun Li, and George R. Uhl. 2015. “Human Cell Adhesion Molecules: Annotated Functional Subtypes and Overrepresentation of Addiction-Associated Genes.” Annals of the New York Academy of Sciences 1349 (1): 83–95. 10.1111/nyas.12776.

Zhou, Hang, Rachel L. Kember, Joseph D. Deak, Heng Xu, Sylvanus Toikumo, Kai Yuan, Penelope A. Lind, et al. 2023. “Multi-Ancestry Study of the Genetics of Problematic Alcohol Use in over 1 Million Individuals.” Nature Medicine 29 (12): 3184–92. 10.1038/s41591-023-02653-5.

Zuk, Or, Eliana Hechter, Shamil R. Sunyaev, and Eric S. Lander. 2012. “The Mystery of Missing Heritability: Genetic Interactions Create Phantom Heritability.” Proceedings of the National Academy of Sciences 109 (4): 1193–98. 10.1073/pnas.1119675109.

