## Supplemental Results and Methods for "EXTENSIVE GENETIC INTERACTIONS (EPISTASIS) LINKED TO ALCOHOL USE DISORDER IN A HIGH-RISK POPULATION"

### Supplementary Materials

#### Supplementary Tables

##### Supplemental Table 1: Demographics of the American Indian cohort.

| Cohort | American Indian (AI) |
| --- | --- |
| Individuals (n) | 742 |
| Family | 170 |
| Gender | M: 317, F: 425 |
| Age^1^ (yrs) | 31.2±13.2 [18, 82] |
| Ethnicity | Native American |

^1^ In the format of mean ± standard deviation [min, max].

##### Supplemental Table 2: Core gene set composed of alcohol use disorder (AUD), alcohol dependence (AD), and molecular target of ethanol associated genes.

ABCB7, ABO, ACE, ACSS3, ADCY1, ADCY10, ADCY2, ADCY3, ADCY4, ADCY5, ADCY6, ADCY7, ADCY8, ADCY9, ADGRL3, ADH1A, ADH1B, ADH1C, ADH4, ADH5, ADH6, ADH7, ADRA2A, AFP, AGO1, AGO2, AKR1A1, AKR1C3, AKR1C4, ALB, ALDH1A1, ALDH1B1, ALDH2, ALDH3B2, ALDH9A1, ALK, ALMS1, ANAPC1, ANKK1, ANKRD7, APOE, AR, ARC, ARID4A, ARSA, ASIC2, ASTN1, ATP12A, ATP4A, AUTS2, AVP, BABRB3, BAG3, BAHCC1, BDNF, BHMT, BRAP, BRD3, C1D, CACNA1C, CACNA1E, CADM1, CADM2, CALCA, CALN1, CAMK2A, CAMK4, CARS1, CARTPT, CASC4, CAT, CBS, CCK, CCKAR, CCKBR, CD4, CDH10, CDH11, CDH12, CDH13, CDH15, CDH18, CDH5, CDH8, CDH9, CDK20, CELF2, CFTR, CHRM2, CHRNA1, CHRNA10, CHRNA2, CHRNA3, CHRNA4, CHRNA5, CHRNA6, CHRNA7, CHRNA9, CHRNB1, CHRNB2, CHRNB3, CHRNB4, CHRND, CHRNE, CHRNG, CLOCK, CNOT4, CNR1, CNRN2, CNTN4, CNTN6, CNTNAP2, COMT, CREB1, CRH, CRHBP, CRHR1, CRHR2, CRP, CSMD1, CSMD3, CTNNA2, CXCL8, CYP2A13, CYP2B6, CYP2E1, CYTL1, DAG1, DBH, DCLK2, DKK2, DPYD, DPYSL2, DRD1, DRD2, DRD3, DRD4, DRD5, DSCAML1, DTNBP1, DUSP8, EBI3, ECHS1, EGF, EGFR, EP300, EPHA8, EPHX1, ERCC1, ERI3, ESR1, F2, FAAH, FABP2, FGAP, FHSB, FKBP5, FOLR1, FSHB, FSTL5, FTO, FUT2, FYN, GABBR1, GABRA1, GABRA2, GABRA3, GABRA4, GABRA5, GABRA6, GABRB1, GABRB2, GABRB3, GABRD, GABRE, GABRG1, GABRG2, GABRG3, GABRP, GABRQ, GABRR1, GABRR2, GABRR3, GAD1, GAD2, GAL, GALR1, GALR2, GALR3, GAP43, GAPDH, GATA4, GCKR, GEMIN4, GGH, GGT1, GH1, GHRL, GHS, GHSR, GINS2, GLI2, GLRA1, GLRA2, GLRA3, GLRB, GLUL, GNB3, GPHN, GPT, GPT2, GRIA1, GRIA2, GRIA3, GRIA4, GRID1, GRID2, GRIK1, GRIK2, GRIK3, GRIK4, GRIK5, GRIN1, GRIN2A, GRIN2B, GRIN2C, GRIN2D, GRIN3A, GRIN3B, GRM1, GRM2, GRM3, GRM4, GRM5, GRM6, GRM7, GRM8, GSTM1, HAMP, HCN1, HCN2, HCN3, HCN4, HCRT, HDAC2, HERPUD1, HFE, HLA-C, HLA-DRA, HMGB1, HNMT, HOMER1, HOMER2, HS6ST3, HTR1A, HTR1B, HTR2A, HTR2C, HTR3A, HTR3B, HTR3C, HTR3D, HTR3E, HTR4, HTR7, IGF2BP1, IL10, IL17A, IL1A, IL1B, IL1R1, IL1RN, IL6, INPP4B, INS, IPO11, ISL1, JUN, KANK1, KCNJ3, KCNJ5, KCNJ6, KCNJ9, KCNK2, KCNMA1, KCNMB1, KCNMB2, KCNMB3, KCNMB4, KCNN1, KCNN2, KCNN3, KCNN4, KCNQ1, KCNQ2, KCNQ3, KCNQ4, KCNQ5, KCTD16, KIAA0040, KLB, KLF11, KPNA3, LEP, LHB, LILRA1, LINC02347, LINC02694, LOC126807122, LRP8, LRRC25, LRRC38, LRRC52, LRRC56, MAOA, MAOB, MAPT, MBP, MCM5, MGLL, MICB, MIR106B, MIR140, MIR146A, MIR181B1, MIR196A1, MIR199A1, MIR21, MIR214, MIR223, MIR382, MIR412, MIR486-1, MIR9-1, MLXIPL, MMP2, MMP9, MOBP, MOG, MPDZ, MTHFR, MYO15A, NAP1L4, NAT1, NCAM1, NEUROD2, NF1, NFKB1, NGF, NKAIN1, NKAIN2, NLGN4X, NMUR2, NPS, NPSR1, NPY, NPY1R, NPY2R, NPY5R, NQO2, NR4A2, NRDC, NRXN3, NTRK2, NTS, NTSR1, OGN, OPRD1, OPRK1, OPRL1, OPRM1, OSBPL5, OSBPL9, OXT, OXTR, PCDH10, PCDH12, PDE10A, PDE4B, PDE4C, PDYN, PECR, PENK, PER3, PHF3, PHLDA2, PIK3R1, PKHD1, PKNOX2, PNOC, PNPLA3, POMC, POR, PPARA, PPARG, PPM1G, PPP1R13B, PPP1R1B, PPP1R3B, PRKCA, PRKCB, PRKCD, PRKCE, PRKCG, PRKCH, PRKCI, PRKCQ, PRKCZ, PRKG2, PRL, PTK2B, PTP4A1, RABGAP1L, RACK1, RASGRF2, RASIP1, RBX1, RCBTB1, REN, RFC1, RFX4, RGS4, ROBO2, RPS6KA4, RSRC1, RUNX1T1, SAT1, SCN8A, SDHAF3, SEMA5A, SERINC2, SGCE, SGIP1, SHBG, SIGMAR1, SIX3, SLC17A5, SLC18A2, SLC19A3, SLC1A2, SLC22A18, SLC25A37, SLC29A1, SLC39A8, SLC46A1, SLC4A8, SLC6A1, SLC6A2, SLC6A3, SLC6A4, SLC6A5, SLC6A9, SLC9A8, SLCO3A1, SNCA, SNORA54, SNRNP70, SPG21, SRD5A1, SRD5A2, ST18, STON2, TAC1, TACR1, TACR3, TAGLN3, TAS2R13, TAS2R16, TAS2R38, TBX19, TBX6, TCF4, TESK2, TF, TFAP2B, TGFB1, TH, THEMIS, THSD7B, TIA1, TIPARP, TKT, TLR4, TNF, TNRC6A, TP53, TPH1, TPH2, TRH, TSNARE1, TSPAN5, TSPO, TTC12, UBAP2, UCN1, UCN2, UCN3, VRK2, VWF, XPO7, XRCC5, ZCCHC14, ZIC4, ZNF536, ZNF699, ZNF804A

##### Supplemental Table 3. Alcohol-related life events in the clinical course of AUD.

The order of the events was based on the mean age of the occurrence with the first event happening earliest and the last (36^th^) event occurring latest in a lifetime [Ehlers et al., 2004]. Severity weights that were assigned to events were used to compute AUD severity level [Peng et al., 2019].

| Severity weight 1 | Severity weight 2 | Severity weight 3 |
| --- | --- | --- |
| 1. Arguments | 13. Binges | 25. Arrested for alcohol related behavior |
| 2. Physical fights | 14. Tolerance | 26. Problems in love relationship |
| 3. Problems at work/school | 15. Interfered with work | 27. Considered excessive drinker |
| 4. Problems with family, friends | 16. Self injury while drunk | 28. Guilt |
| 5. Hitting others without fighting | 17. Decreased important activities | 29. Wanted to quit 3+ times |
| 6. Objections from family, friends | 18. Inability to change drinking behavior | 30. Withdrawal |
| 7. Drank while in hazardous situations | 19. Morning drinking | 31. Unable to quit/cut down |
| 8. Drank when not intended | 20. Drank more than intended | 32. Arrested for DUI |
| 9. Lost friends | 21. Used rules for drinking | 33. Shakes |
| 10. Blackouts | 22. Little time for non-drinking activities | 34. Continues despite health problems |
| 11. Hit/threw things | 23. Strong desire for alcohol | 35. Health problems occurred |
| 12. Hit family member | 24. Psychological impairment | 36. Sought professional help |

Supplemental Table 4: Table of interacting gene and/or regulatory element pairs.

Interacting elements for which there is prior evidence of interaction are bolded. The p-values provided were obtained post bi-clustering using hypergeometric distribution. Genes with existing association to AUD/AD are underlined. After each regulatory element, in parenthesis, listed are protein coding genes regulated by it, with GeneHancer score of > 25. Note that some gene and/or regulatory element pairs may share p-values, as they came from the same interval pair and p-values were generated per interval pair. The genes and/or regulatory elements listed using ‘/’ share SNPs within same interval.

| Interacting Element #1 | Interacting Element #2 | # of interacting SNP pairs | Prior Evidence of Interaction | p-value(s) |
| --- | --- | --- | --- | --- |
| **ABAT** | **PRKCB,** GH16J023835 (**PRKCB**) | 14, 5 | Both are present together in multiple neuronal pathways. | 1.35e-112 |
| ACOT12 | MAML2 | 6 |  | 6.89e-41 |
| ACOX3 | CELF2 | 2 |  | 3.26e-17 |
| ADGRL3 | APPBP2 | 6 |  | 1.53e-40 |
| ALK | NAT1 | 4 |  | 4.05e-24 |
| APP | SCN2B | 19 |  | 6.33e-82 |
| **ASIC2** | **KIF5B**, PRKCQ | 4, 10 | Both ASIC2 and KIF5B may interact with Syntabulin. [[5](10.1038/srep20924)] | 2.23e-26, 1.10e-57 |
| ASTN1 | IL1RN | 5 |  | 9.10e-29 |
| BRD7 | CTNNA1 | 8 |  | 4.70e-45 |
| **CACNA2D1** | **IL1A** | 6 | Both are present together in MAPK pathway. | 1.92e-43 |
| CALCR | DCAF11 | 3 |  | 6.90e-25 |
| CARD11 | RUNX1 | 5 |  | 3.30e-26 |
| CDH13 | GREB1, KCNK2, PRKN, RFC1, SLC25A37 | 13,4,3,5,2 |  | 2.92e-72, 3.62e-30, 2.09e-17, 1.26e-29, 1.28e-16 |
| CGA | ZFPM2 | 7 |  | 7.50e-50 |
| **CHRM2** | **GRM4** | 3 | Present together in various signaling and binding pathways. | 4.25e-25 |
| CHRNG | INSR | 11 |  | 7.11e-84 |
| **CNTNAP2** | INPP4B, GABRR3, GRM8, **WRN** | 52, 2, 15, 5 | Present together in ovarian insufficiency pathway. | <2.22e-308, 9.22e-17, 3.64e-66, 1.00e-33 |
| CNTN2 | HS6ST3 | 3 |  | 8.07e-20 |
| CNTN6 | SGCD | 4 |  | 2.24e-28 |
| **CSMD1** | **ERBB4**, CAMK2A, **DLGAP1,** MAML2, NLGN1 | 3,3,3,2,25 | CSMD1 and ERBB4 may interact directly [[2](https://doi.org/10.1186/s13046-021-02042-1)]. CSMD1 and DLGAP1 have a StringDB connection. | 1.38e-22, 1.59e-24, 6.39e-23, 2.19e-18, 5.26e-161 |
| **CTBP2** | **KCNQ4** | 2 | Present together in Reactome sensory pathway. | 1.17e-16 |
| **DLG2** | KAT2B, **PRKCA** | 2, 11 | Present together in multiple neuronal pathways. | 9.32e-17, 1.10e-76 |
| DPYD | SNRPG, TIA1 | 3, 35 |  | 1.17e-18, 1.10e-138 |
| DSCAML1 | F2R | 5 |  | 1.54e-34 |
| EIF4E | SIN3A | 29 |  | 2.10e-136 |
| EP400 | NKAIN2 | 6 |  | 5.68e-45 |
| ERCC6 | CDH13 / GH16J082626 (CDH13) | 3 |  | 1.71e-21 |
| **EXOSC7** | **TFAP2A** | 3 | Present together in ciliary landscape pathway. | 1.67e-24 |
| FBXL7 | SLIT2 | 3 |  | 7.68e-24 |
| FGF6 | SLC6A4 | 2 |  | 6.27e-17 |
| FSTL5 | PPARGC1A | 40 |  | 1.44e-264 |
| FTO | FTO | 4 |  | 3.30e-24 |
| **GABRB3** | **KCNN4 /** GH19J043773 (**KCNN4,** ZNF45, ZNF180, ZNF225, ZNF224, ZNF284, ZNF227, ZNF223, ZNF283, ZNF234, ZNF235, XRCC1, ZNF226, ZNF221, ZNF576, ZNF230, ZNF229, PINLYP, SMG9, CEACAM19, ZNF285) | 54 | Both GABRB3 and KCNN4 are present together in multiple neuronal system pathways. | 1.34e-199 |
| GABRG3 | APP, CADM2, MBD2 | 20, 8, 5 |  | 2.21e-54, 9.54e-43, 3.61-37 |
| GNGT1 | PKNOX2 | 2 |  | 7.84e-18 |
| **GRIN2B** | **PDE10A,** PRKCH | 5, 7 | GRIN2B and PDE10A are present together in a neuronal pathway. | 6.22e-29, 3.20e-45 |
| GUCY1A2 | GABBR1 / HLA-F | 67 |  | <2.22e-308 |
| HLA-DRB1 | GRK2, PPP1CA | 3, 11 |  | 3.54e-69, 3.54e-69 |
| HLA-F | PAK5 | 8 |  | 3.92e-49 |
| **HLA-G** | PAK5, CELF2, EXOSC10, **MTOR** | 8,6,19,73 | Both HLA-G and MTOR are present in the Reactome adaptive immune system pathway. | 3.92e-49, 1.20e-47, 1.40e-86, 7.30e-252 |
| **HSPD1** | **HLA-C / HLA-B /** MICB | 5 | HSPD1 and HLA-B/HLA-C are present together in Kegg diabetes pathway. | 4.78e-84 |
| HSD17B4 | ITGB8 | 4 |  | 5.89e-27 |
| **HTR2A** | **IL6R** | 3 | Both are present in sudden infant death syndrome pathway. | 1.00e-20 |
| KCNQ3 | JAK2, KCTD1 | 16, 12 |  | 5.68e-114, 3.65e-82 |
| LUC7L | UBA52 | 7 |  | 1.78e-50 |
| **MAGI1** | CSMD1, NLRP3, **PPP2R2C,** PLXNA4 | 4, 2, 6, 7 | Present together in Kegg tight junction pathway. | 1.08e-27, 2.21e-17, 2.63e-44, 1.96e-48 |
| **MAPK9** | **TIAM1** | 20 | Present together in neurotrophic, signaling, and other pathways. | 7.48e-138 |
| **MAPT** | LRP2, **GNG7** | 3, 26 | Both MAPT and GNG7 are present together in multiple neuronal pathways. | 4.78e-23, 8.33e-155 |
| **MTA3** | **CDH2**, CDH13, HDAC9 | 8, 3,10 | MTA3 may impact expression levels of CDH2 and interact in context of some cancers. [[4](https://doi.org/10.3390/cancers13236098)] | 8.08e-55, 3.08e-23, 9.76e-34 |
| **MYCN** | **LDHA** | 6 | May interact in context of certain cancers. [[3](https://doi.org/10.1158/1078-0432.CCR-17-2578)] | 5.71e-44 |
| NLRP12 | KSR2 | 2 |  | 1.95e-16 |
| NPAS2 | HCN1 | 47 |  | 2.18e-205 |
| **NRXN1** | **CTNNA2,** YES1 | 3, 5 | NRXN1 and CTNNA2 are connected according to StringDB. | 8.59e-25, 2.58e-26 |
| OPRM1 | SNRPN | 5 |  | 1.72e-37 |
| **PIK3R1,** GH05J068214 (**PIK3R1**) | **MICA / MICB** | 3, 1 | PIK3R1 and MICA/MICB are present together in neuronal and immune system pathways. | 8.49e-29 |
| PLXNA2 | KLB, RFC1 | 10, 6 |  | 1.59e-66, 5.08e-40 |
| PPP3CA | POMT1 | 35 |  | 1.44e-237 |
| RABGAP1L | HLA-G / HLA-A | 3 |  | 6.24e-19 |
| RASGRF1 | HADH, LEF1 | 44, 16 |  | <2.22e-308 |
| ROBO2 | KCNQ5, KLHL3 | 4, 2 |  | 2.26e-29, 1.53e-16 |
| **RYR2** | **ADCY9,** GH16J004110 (**ADCY9**) | 3 | Both are present together in calcium, heart, and other pathways. | 7.69e-23 |
| **SF3B1** | **HLA-C / HLA-B / MICB** | 8 | There is some limited evidence of HLA-B and SF3B1 interacting [[1](https://ash.confex.com/ash/2020/webprogram/Paper139907.html)]. SF3B1, MICB, and HLA-C are present together in an IL-24 signaling pathway. | 4.78e-84 |
| SGCZ | MMP9, GH20J046007 (MMP9) | 2,1 |  | 8.20e-24 |
| SLC17A5 | TERF1 | 8 |  | 1.47e-44 |
| SLIT3 | TAB2 | 9 |  | 1.31e-58 |
| SUMF1 | PPP2R2B, GRB2 | 8, 27 |  | 3.52e-53, 3.15e-117 |
| **TCF7L1** | **EDNRB** | 24 | Present together in Kegg melanogenesis pathway. | 1.16e-143 |
| VAV3 | SLC17A5 | 14 |  | 4.53e-86 |

Supplemental Table 5: Enrichment analysis of interacting genes using Enrichr and FUMA GWAS.

Pathways identified by both Enrichr and FUMA GWAS were only listed once.

| Name | p-value | Adjusted p-value | Database |
| --- | --- | --- | --- |
| Endosomal/vacuolar pathway | 4.13e-5 | 1.71e-2 | Enrichr BioPlanet 2019 |
| Cell adhesion molecules (CAMs) | 4.14e-5 | 1.71e-2 |  |
| Autoimmune thyroid disease | 2.72e-4 | 2.40e-2 | Enrichr KEGG 2021 Human |
| Type I diabetes mellitus | 3.00e-4 | 2.40e-2 |  |
| Graft versus host disease | 1.76e-3 | 9.24e-2 |  |
| Allograft rejection | 1.93e-3 | 9.24e-2 |  |
| Antigen Processing And Presentation Of Endogenous Peptide Antigen Via MHC Class I Via ER Pathway | 8.80e-8 | 4.29e-5 | Enrichr GO Biological Process 2023 |
| Antigen Processing And Presentation Of Endogenous Peptide Antigen Via MHC Class I Via ER Pathway, TAP-independent | 8.80e-8 | 4.29e-5 |  |
| Antigen Processing And Presentation Of Endogenous Peptide Antigen Via MHC Class Ib | 8.80e-8 | 4.29e-5 |  |
| Antigen Processing And Presentation Of Peptide Antigen Via MHC Class Ib | 8.80e-8 | 4.29e-5 |  |
| Antigen Processing And Presentation Of Endogenous Peptide Antigen | 3.21e-7 | 1.25e-4 |  |
| Positive Regulation Of T Cell Mediated Cytotoxicity | 1.71e-6 | 5.56e-4 |  |
| Positive Regulation Of T Cell Mediated Immunity | 4.90e-6 | 1.37e-3 |  |
| Positive Regulation Of Leukocyte Mediated Cytotoxicity | 1.92e-5 | 4.67e-3 |  |
| Regulation Of T Cell Mediated Cytotoxicity | 3.10e-5 | 6.72e-3 |  |
| Homophilic Cell Adhesion Via Plasma Membrane Adhesion Molecules | 7.49e-5 | 1.46e-2 |  |
| Cell-Cell Adhesion Via Plasma-Membrane Adhesion Molecules | 2.16e-4 | 3.84e-2 |  |
| Positive Regulation Of Axonogenesis | 2.92e-4 | 4.75e-2 |  |
| Neuron Projection | 1.12e-4 | 2.15e-2 | Enrichr GO Cellular Component 2023 |
| Axon | 2.34e-4 | 2.24e-2 |  |
| Phagocytic Vesicle Membrane | 4.11e-4 | 2.27e-2 |  |
| Lumenal Side of Endoplasmic Reticulum Membrane | 6.87e-4 | 2.27e-2 |  |
| Recycling Endosome | 6.87e-4 | 2.27e-2 |  |
| Recycling Endosome Membrane | 7.09e-4 | 2.27e-2 |  |
| Phagocytic Vesicle | 1.93e-3 | 4.62e-2 |  |
| Early Endosome | 1.93e-3 | 4.62e-2 |  |
| BENPORATH ES with H3K27ME3 | 6.43e-6 | 4.15e-2 | FUMA Curated Gene Sets |
| Stein estrogen response not via ESRRA | 1.28e-5 | 4.15e-2 |  |
| Midbrain human GABAergic neurons | 1.41e-8 | 1.17e-5 | FUMA Cell type signatures |
| Midbrain human GABAergic neuroblasts | 2.53e-7 | 1.05e-4 |  |
| Fetal lung visceral neurons | 2.06e-4 | 1.92e-2 |  |
| Brain frontal cortex | 2.51e-7 | NA | FUMA Tissue Specificity (Upregulated) |
| Brain anterior cingulate cortex | 3.98e-7 | NA |  |
| Brain caudate basal ganglia | 1.00e-6 | NA |  |
| Brain nucleus accumbens basal ganglia | 1.99e-6 | NA |  |
| Brain hypothalamus | 2.50e-5 | NA |  |
| Brain hippocampus | 2.82e-5 | NA |  |
| Brain cortex | 3.02e-5 | NA |  |
| Brain putamen basal ganglia | 3.16e-5 | NA |  |
| Brain cerebellum | 7.94e-4 | NA |  |
| Brain amygdala | 8.32e-4 | NA |  |
| Brain substantia nigra | 1.26e-3 | NA |  |
| Vagina | 1.00e-4 | NA | FUMA Tissue Specificity (Downregulated) |
| Adult body size | 2.05e-7 | 9.05e-4 | FUMA catalog reported genes |
| Body size at age 10 | 8.09e-6 | 1.69e-2 |  |
| Atopic dermatitis | 1.15e-5 | 1.69e-2 |  |
| Adolescent idiopathic scoliosis | 4.20e-5 | 4.29e-2 |  |
| Height | 4.85e-5 | 4.29e-2 |  |

Supplemental Table 6: Enrichment analysis of genes regulated by interacting regulatory elements using Enrichr and FUMA GWAS.

Duplicate pathways were removed. Used GeneHancer threshold score of 25 to select the regulated elements.

| Name | p-value | Adjusted p-value | Database |
| --- | --- | --- | --- |
| Generic Transcription Pathway | 8.26e-9 | 1.87e-6 | Enrichr Reactome 2022 |
| RNA Polymerase II Transcription | 2.46e-8 | 2.80e-6 |  |
| Gene Expression (Transcription) | 7.42e-8 | 5.61e-6 |  |
| Phospholipase C signaling pathway | 7.27e-5 | 6.58e-3 | Enrichr Bioplanet 2019 |
| LPA receptor mediated events | 8.07e-5 | 6.58e-3 |  |
| TrkA receptor signaling pathway | 1.69e-4 | 1.10e-2 |  |
| Salivary secretion | 2.06e-4 | 1.12e-2 |  |
| CREB transcription factor and its extracellular signals | 4.04e-4 | 1.52e-2 |  |
| CXCR4 signaling pathway | 4.48e-4 | 1.52e-2 |  |
| Leukocyte transendothelial migration | 4.60e-4 | 1.52e-2 |  |
| TPO signaling pathway | 4.79e-4 | 1.52e-2 |  |
| VEGFR1 pathway | 5.59e-4 | 1.52e-2 |  |
| eIF4E and p70 S6 kinase regulation | 5.59e-4 | 1.52e-2 |  |
| PDGFA signaling pathway | 6.91e-4 | 1.72e-2 |  |
| Alpha-V beta-3 integrin/OPN pathway | 7.38e-4 | 1.72e-2 |  |
| Phospholipids as signaling intermediaries | 9.96e-4 | 2.10e-2 |  |
| PAC1 receptor pathway | 1.23e-3 | 2.10e-2 |  |
| Plasma membrane estrogen receptor signaling | 1.29e-3 | 2.10e-2 |  |
| Bioactive peptide-induced signaling pathway | 1.29e-3 | 2.10e-2 |  |
| Fc epsilon receptor I signaling in mast cells | 1.29e-3 | 2.10e-2 |  |
| Aldosterone-regulated sodium reabsorption | 1.35e-3 | 2.10e-2 |  |
| VEGF, hypoxia, and angiogenesis | 1.35e-3 | 2.10e-2 |  |
| Growth hormone receptor signaling | 1.42e-3 | 2.10e-2 |  |
| G alpha (z) signaling events | 1.55e-3 | 2.20e-2 |  |
| Chemokine signaling pathway | 1.84e-3 | 2.50e-2 |  |
| Thrombin signaling through protease-activated receptors | 2.15e-3 | 2.60e-2 |  |
| PI3K pathway | 2.15e-3 | 2.60e-2 |  |
| Kit receptor signaling pathway | 2.23e-3 | 2.60e-2 |  |
| Non-small cell lung cancer | 2.23e-3 | 2.60e-2 |  |
| Vibrio cholerae infection | 2.31e-3 | 2.60e-2 |  |
| Mechanism of gene regulation by peroxisome proliferators via PPAR-alpha | 2.48e-3 | 2.70e-2 |  |
| FGF signaling pathway | 2.84e-3 | 2.98e-2 |  |
| Endothelins | 3.12e-3 | 3.17e-2 |  |
| Glioma | 3.21e-3 | 3.17e-2 |  |
| Signaling events mediated by VEGFR1 and VEGFR2 | 3.72e-3 | 3.56e-2 |  |
| Gastric acid secretion | 4.14e-3 | 3.86e-2 |  |
| VEGF signaling pathway | 4.37e-3 | 3.94e-2 |  |
| Phosphatidylinositol signaling system | 4.59e-3 | 3.94e-2 |  |
| Signaling by SCF-KIT | 4.59e-3 | 3.94e-2 |  |
| Fc epsilon receptor I signaling pathway | 4.71e-3 | 3.94e-2 |  |
| MicroRNAs in cardiomyocyte hypertrophy | 5.43e-3 | 4.42e-2 |  |
| Progesterone-mediated oocyte maturation | 5.55e-3 | 4.42e-2 |  |
| Neuronal system | 5.73e-3 | 4.42e-2 |  |
| Gap junction pathway | 6.07e-3 | 4.48e-2 |  |
| Phospholipase C delta-1 interactions in phospholipid-associated cell signaling | 6.48e-3 | 4.48e-2 |  |
| Ion channel and phorbal esters signaling pathway | 6.48e-3 | 4.48e-2 |  |
| Disinhibition of SNARE formation | 6.48e-3 | 4.48e-2 |  |
| ERBB signaling pathway | 6.60e-3 | 4.48e-2 |  |
| Fc gamma receptor-mediated phagocytosis | 6.60e-3 | 4.48e-2 |  |
| G-protein signaling pathways | 6.74e-3 | 4.48e-2 |  |
| Pancreatic secretion | 7.58e-3 | 4.75e-2 |  |
| GnRH signaling pathway | 7.58e-3 | 4.75e-2 |  |
| Melanogenesis | 7.58e-3 | 4.75e-2 |  |
| Signaling by ERBB2 | 7.73e-3 | 4.75e-2 |  |
| Amoebiasis | 8.17e-3 | 4.84e-2 |  |
| Fibroblast growth factor receptor pathway | 8.17e-3 | 4.84e-2 |  |
| Pathways in cancer | 8.38e-3 | 4.86e-2 |  |
| G alpha i pathway | 8.63e-3 | 4.86e-2 |  |
| Gamma-secretase-mediated ErbB4 signaling pathway | 9.07e-3 | 4.86e-2 |  |
| Activation of PKC through G-protein coupled receptors | 9.07e-3 | 4.86e-2 |  |
| Signaling by EGFR in cancer | 9.09e-3 | 4.86e-2 |  |
| Epidermal growth factor receptor (EGFR) pathway | 9.09e-3 | 4.86e-2 |  |
| Macrophage Stimulating Protein MSP Signaling | 3.64e-4 | 2.51e-2 | Enrichr WikiPathways 2023 |
| PDGFR Beta Pathway | 6.46e-4 | 2.51e-2 |  |
| GPR143 In Melanocytes And Retinal Pigment Epithelium Cells | 6.91e-4 | 2.51e-2 |  |
| Hepatitis B Infection | 9.66e-4 | 2.51e-2 |  |
| Kisspeptin Receptor System In The Ovary | 1.17e-3 | 2.51e-2 |  |
| Bladder Cancer | 1.23e-3 | 2.51e-2 |  |
| Chronic Hyperglycemia Impairment Of Neuron Function | 1.62e-3 | 2.86e-2 |  |
| Oncostatin M Signaling Pathway | 2.40e-3 | 3.07e-2 |  |
| IL 18 Signaling Pathway | 2.40e-3 | 3.07e-2 |  |
| AGE RAGE Pathway | 3.31e-3 | 3.89e-2 |  |
| Glioblastoma Signaling Pathways | 5.06e-3 | 4.25e-2 |  |
| EGFR Tyrosine Kinase Inhibitor Resistance | 5.06e-3 | 4.25e-2 |  |
| Acute Viral Myocarditis | 5.43e-3 | 4.25e-2 |  |
| Orexin Receptor Pathway | 5.81e-3 | 4.25e-2 |  |
| B Cell Receptor Signaling Pathway | 6.33e-3 | 4.16e-2 |  |
| Hippo Signaling Regulation Pathways | 7.15e-3 | 4.58e-2 |  |
| Metastatic Brain Tumor | 7.78e-3 | 4.77e-2 |  |
| Herpes simplex virus 1 infection | 7.26e-20 | 1.06e-17 | Enrichr Kegg 2021 |
| GnRH secretion | 7.71e-5 | 5.63e-3 |  |
| Insulin secretion | 1.86e-4 | 7.98e-3 |  |
| Inflammatory mediator regulation of TRP channels | 2.73e-4 | 7.98e-3 |  |
| Cholinergic synapse | 4.15e-4 | 8.82e-3 |  |
| Growth hormone synthesis, secretion and action | 4.83e-4 | 8.82e-3 |  |
| Relaxin signaling pathway | 6.11e-4 | 9.92e-3 |  |
| Estrogen signaling pathway | 7.28e-4 | 1.06e-2 |  |
| Carbohydrate digestion and absorption | 1.69e-3 | 1.88e-2 |  |
| Endocrine and other factor-regulated calcium reabsorption | 2.15e-3 | 1.88e-2 |  |
| Diabetic cardiomyopathy | 2.25e-3 | 1.88e-2 |  |
| Regulation of lipolysis in adipocytes | 2.31e-3 | 1.88e-2 |  |
| Proteoglycans in cancer | 2.32e-3 | 1.88e-2 |  |
| Rap1 signaling pathway | 2.48e-3 | 1.91e-2 |  |
| Human cytomegalovirus infection | 3.02e-3 | 2.10e-2 |  |
| Chemical carcinogenesis | 3.58e-3 | 2.37e-2 |  |
| Thyroid hormone synthesis | 4.25e-3 | 2.45e-2 |  |
| GABAergic synapse | 5.94e-3 | 2.45e-2 |  |
| Morphine addiction | 6.20e-3 | 2.45e-2 |  |
| Circadian entrainment | 7.01e-3 | 2.45e-2 |  |
| Prostate cancer | 7.01e-3 | 2.45e-2 |  |
| Aldosterone synthesis and secretion | 7.15e-3 | 2.45e-2 |  |
| Choline metabolism in cancer | 7.15e-3 | 2.45e-2 |  |
| MicroRNAs in cancer | 7.36e-3 | 2.45e-2 |  |
| Longevity regulating pathway | 7.73e-3 | 2.45e-2 |  |
| Parathyroid hormone synthesis, secretion and action | 8.32e-3 | 2.59e-2 |  |
| Insulin resistance | 8.63e-3 | 2.62e-2 |  |
| HIF-1 signaling pathway | 8.78e-3 | 2.62e-2 |  |
| TNF signaling pathway | 9.25e-3 | 2.70e-2 |  |
| Glutamatergic synapse | 9.57e-3 | 2.74e-2 |  |
| Sphingolipid signaling pathway | 1.04e-2 | 2.92e-2 |  |
| Thyroid hormone signaling pathway | 1.07e-2 | 2.96e-2 |  |
| Platelet activation | 1.12e-2 | 3.04e-2 |  |
| Natural killer cell mediated cytotoxicity | 1.25e-2 | 3.31e-2 |  |
| Vascular smooth muscle contraction | 1.28e-2 | 3.35e-2 |  |
| Fluid shear stress and atherosclerosis | 1.40e-2 | 3.58e-2 |  |
| Spinocerebellar ataxia | 1.47e-2 | 3.71e-2 |  |
| Phospholipase D signaling pathway | 1.57e-2 | 3.83e-2 |  |
| Retrograde endocannabinoid signaling | 1.57e-2 | 3.83e-2 |  |
| mTOR signaling pathway | 1.70e-2 | 3.99e-2 |  |
| Oxytocin signaling pathway | 1.70e-2 | 3.99e-2 |  |
| Hepatocellular carcinoma | 2.00e-2 | 4.63e-2 |  |
| Influenza A | 2.09e-2 | 4.76e-2 |  |
| Cis-Regulatory Region Sequence-Specific DNA Binding | 3.86e-4 | 1.03e-2 | Enrichr GO Molecular Function 2023 |
| RNA Polymerase II Cis-Regulatory Region Sequence-Specific DNA Binding | 4.40e-4 | 1.03e-2 |  |
| RNA Polymerase II Transcription Regulatory Region Sequence-Specific DNA Binding | 7.46e-4 | 1.17e-2 |  |
| DNA-binding Transcription Activator Activity, RNA Polymerase II-specific | 9.95e-4 | 1.17e-2 |  |
| Neurotrophin TRK Receptor Binding | 6.48e-3 | 4.35e-2 |  |
| Neurotrophin TRKA Receptor Binding | 6.48e-3 | 4.35e-2 |  |
| ADP-D-ribose Modification-Dependent Protein Binding | 6.48e-3 | 4.35e-2 |  |
| Oxidized DNA Binding | 9.07e-3 | 4.57e-2 |  |
| Phosphatase Binding | 9.25e-3 | 4.57e-2 |  |
| Protein Phosphatase Binding | 9.73e-3 | 4.57e-2 |  |
| Histone H3 Kinase Activity | 1.16e-2 | 4.97e-2 |  |
| KOYAMA SEMA3B Targets Up | 6.59e-6 | 2.14e-2 | FUMA Curated Gene Sets |
| [BIOCARTA PLC p](http://www.gsea-msigdb.org/gsea/msigdb/human/geneset/BIOCARTA_PLC_PATHWAY)athway | 3.94e-5 | 4.26e-2 | FUMA All Canonical Pathways |
| [PID Lysophospholipid pathway](http://www.gsea-msigdb.org/gsea/msigdb/human/geneset/PID_LYSOPHOSPHOLIPID_PATHWAY) | 4.14e-5 | 4.26e-2 |  |
| [PAR1 pathway](http://www.gsea-msigdb.org/gsea/msigdb/human/geneset/BIOCARTA_PAR1_PATHWAY) | 1.67e-4 | 9.95e-3 | FUMA Biocarta |
| [EGF pathway](http://www.gsea-msigdb.org/gsea/msigdb/human/geneset/BIOCARTA_EGF_PATHWAY) | 3.80e-4 | 9.95e-3 |  |
| [GH pathway](http://www.gsea-msigdb.org/gsea/msigdb/human/geneset/BIOCARTA_GH_PATHWAY) | 3.80e-4 | 9.95e-3 |  |
| [FCER1 pathway](http://www.gsea-msigdb.org/gsea/msigdb/human/geneset/BIOCARTA_FCER1_PATHWAY) | 7.16e-4 | 1.61e-2 |  |
| [TCR pathway](http://www.gsea-msigdb.org/gsea/msigdb/human/geneset/BIOCARTA_TCR_PATHWAY) | 9.23e-4 | 1.93e-2 |  |
| DNA binding transcription factor activity | 2.09e-5 | 1.88e-2 | FUMA GO Molecular Function |
| Transcription regulatory activity | 3.19e-5 | 1.88e-2 |  |
| Ovary | 3.98e-5 | NA | FUMA Tissue Specificity |
| Prostate | 4.47e-5 | NA |  |
| Thyroid | 7.84e-5 | NA |  |
| Heart Atrial Appendage | 1.99e-4 | NA |  |
| Uterus | 2.24e-4 | NA |  |

#### Supplementary Figures

##### Supplemental Figure 1: AUD severity score is highly aligned and correlated with DSM5 AUD diagnosis in AI.

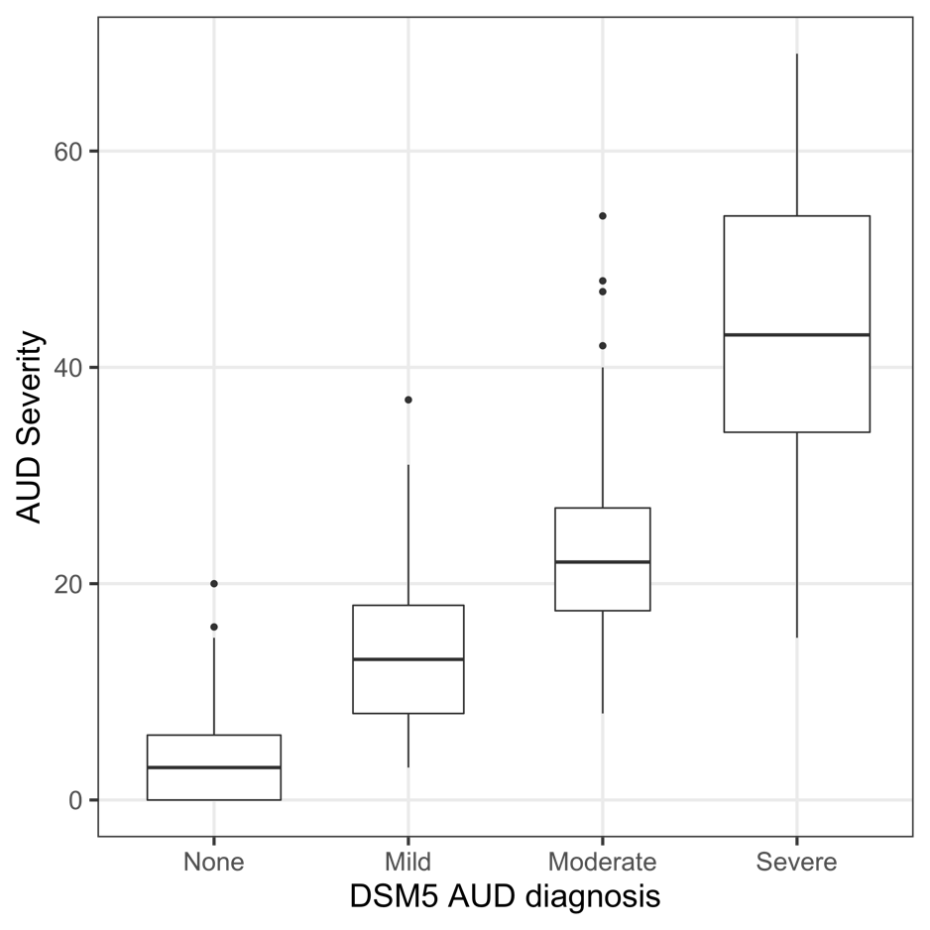

Spearman’s rank correlation = 0.91

Pearson’s correlation = 0.87

##### Supplemental Figure 2: Tissue specificity analysis (upregulated) of the interacting genes using expanded core gene set as background in FUMA.

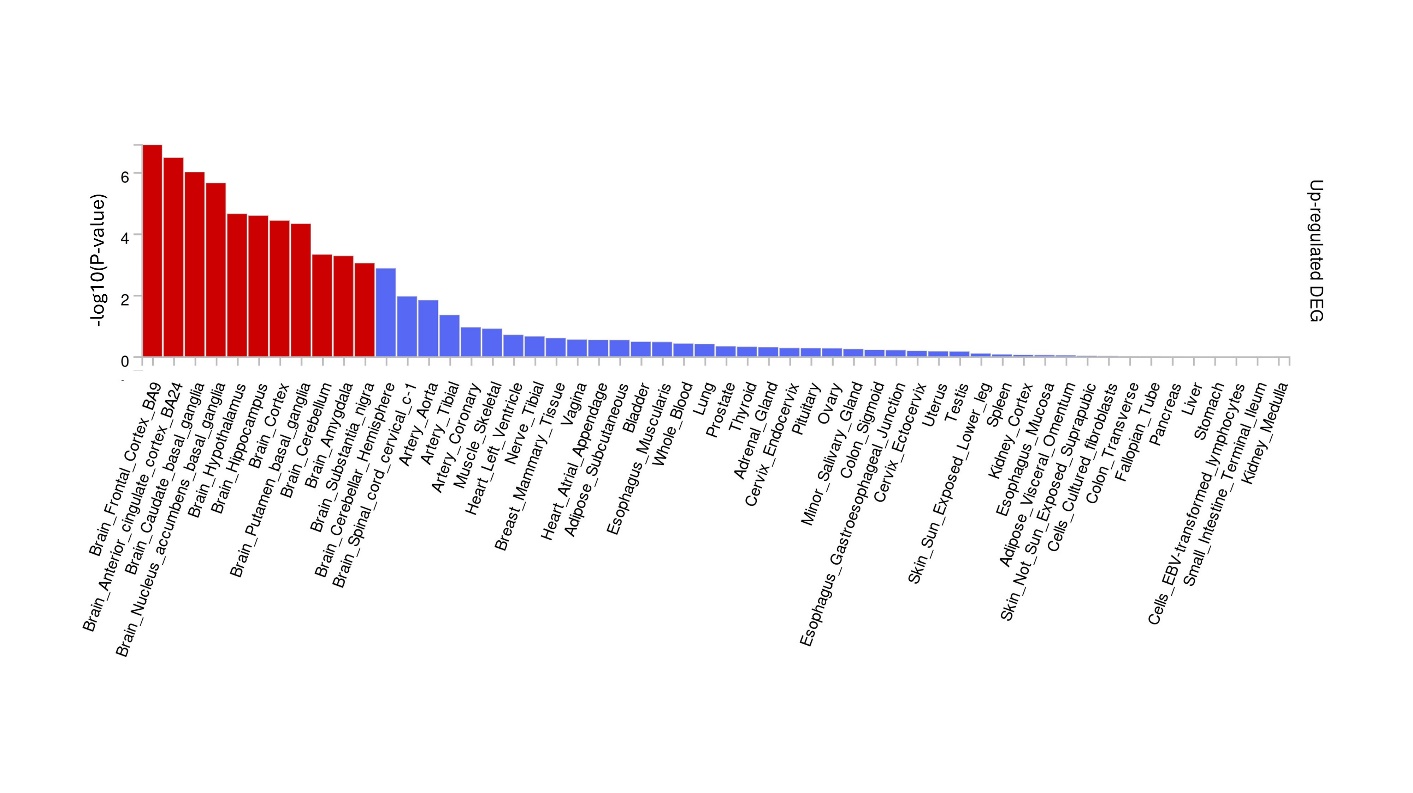

##### Supplemental Figure 3: Cell type enrichment analysis of interacting genes using expanded core gene set as background.

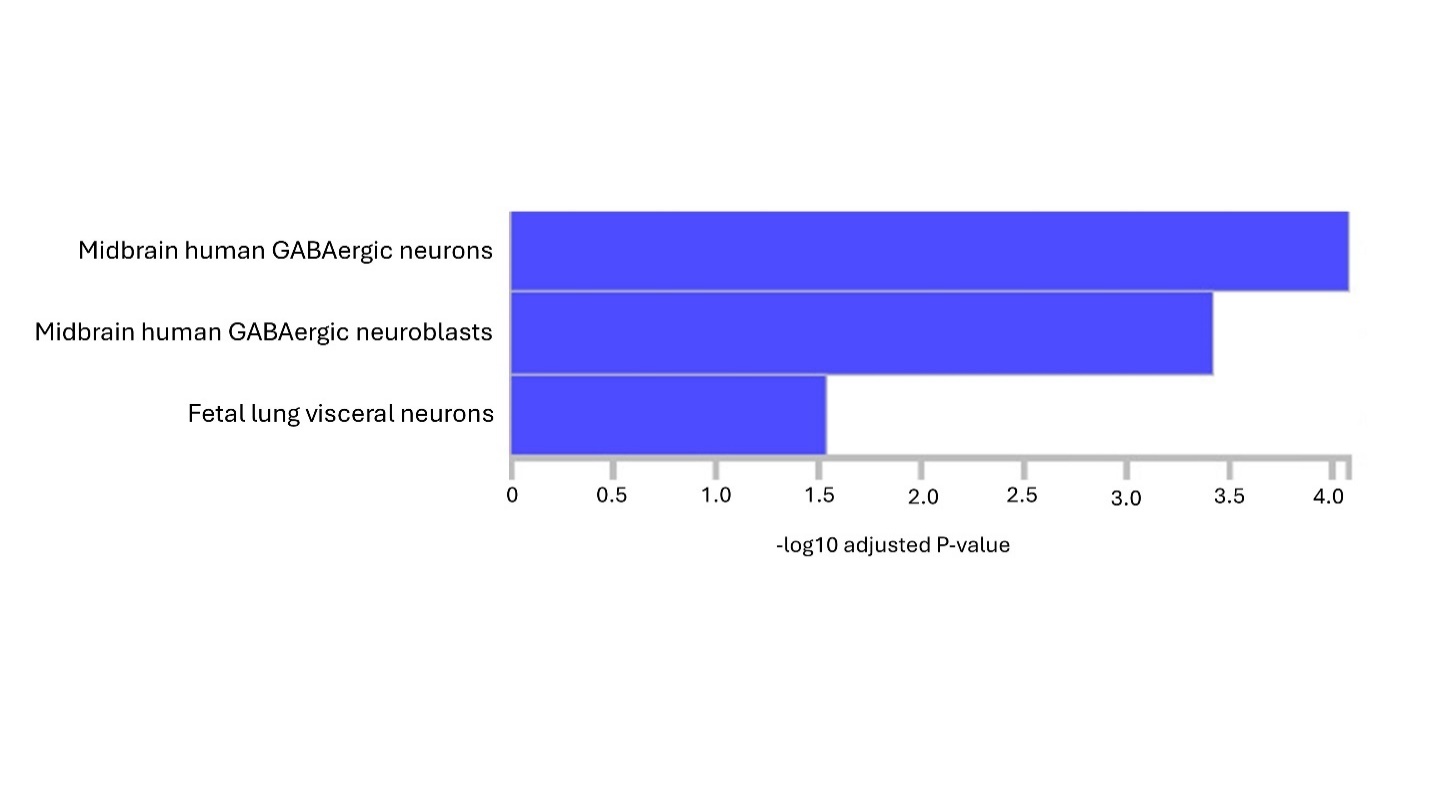

#### Supplementary Methods

##### S1. Participants

American Indian participants were recruited from eight geographically contiguous reservations with a total population of about 3,000 individuals. To be included in the study, participants had to be between the ages of 18 and 70 years, and mobile enough to be transported from their home to The Scripps Research Institute (TSRI). Participants were recruited using a combination of a venue-based method for sampling hard-to-reach populations (Kalton and Anderson, 1986; Muhib et al., 2001) and a respondent-driven procedure (Heckathorn, 1997) that has been described elsewhere (Ehlers et al., 2004). Approximately half of the participants were recruited using each method. A 10-25% rate of refusal using the venue method occurred depending on venue. Refusal rates were higher at tribal libraries and stores than health clinics and tribal halls or culture centers. The refusal rate in the respondent-driven procedure is not known. The protocol for the study was approved by the Institutional Review Board (IRB) of TSRI, and the board of the Indian Health Council, a tribal review group overseeing health issues for the reservations where the recruitment was undertaken. Written informed consent was obtained from each participant after the study was fully explained.

##### S2. AUD severity phenotype

The SSAGA is a semi-structured, poly-diagnostic psychiatric interview that has undergone both reliability and validity testing based on DSM-IV (Bucholz et al., 1994; Hesselbrock et al., 1999). It has also been used in another AI sample (Hesselbrock et al., 2003, 2000), which collected information including demographics, psychiatric history, and symptoms of substance use disorders. Diagnoses of lifetime DSM-5 AUD (mild, moderate, or severe) were generated using the SSAGA. A research psychiatrist/addiction specialist made all best final diagnoses of AUD (Ehlers et al., 2004; Gilder et al., 2004). In addition, the interview retrospectively asks about the occurrence of alcohol-related life events, and the age at which the problem first occurred, from which one of the main quantitative phenotypes for this study, the severity level of AUD, was derived.

Schuckit and colleagues (Schuckit et al., 1993) described measures of the clinical course of alcoholism based on the relative order of the appearance of major “alcohol-related life events”. The clinical course of AUD for the American Indian cohort was previously described (Ehlers et al., 2015, 2004). The severity level of AUD was indexed by 36 alcohol-related life events (Table S3) in the clinical course of the disorder, with life events given a severity weight of 1 for events 1-12; 2 for 13-24; and 3 for 25-36. AUD severity was then calculated as the sum of the severity weights of the 36 life events (Peng et al., 2019).

##### S3. Genotyping of American Indian cohort

The whole genome sequencing pipeline of the AI cohort was previously described (Bizon et al., 2014). In brief: blood derived DNA was sequenced using Illumina low-coverage whole genome sequencing (LCWGS): as well as genotyped using an Affymetrix Exome1A chip. The pair-end sequencing was performed on HiSeq2000 sequencers (Illumina). About 80% of the samples had coverage between 3X and 12X: approximately evenly distributed. Reads from whole genome sequencing were aligned to the GRCh37/hg19 human reference genome using BMA: and realigned near indels with GATK (DePristo et al., 2011). Variants were called using both GATK Unified Genotyper following the best practices for low-coverage samples (Van der Auwera et al., 2013) and the LD-aware variant caller Thunder (Li et al., 2011). Imputation was carried out using the program Thunder. Qualities of variant calling were assessed through a comparison between the sequencing results to genotypes generated on the exome array for the same set of subjects. The assessment showed nearly all of common variants and a high percentage of rare variants in the samples were correctly called. The median concordance rate was 97.5%. The median false positive rate was 0.3% (Bizon et al., 2014). The variant calling process resulted 23,550,342 genome-wide variants for 750 AI individuals.

Further quality control (QC) and filtering were performed as follows prior to genome-wide association analyses: removing variants with high missing rate (>5%), variants out of Hardy-Weinberg equilibrium (HWE) (*p*<1E-6), variants having high Mendel error rate (>5%), and removing individuals if missing >2% genotypes.

##### S4. Core Gene Set

Sources Used:

- Diseases 2.0 Database (Jensen Lab) (Grissa et al., 2022) – last updated on 04/26/2024.
- Kegg Disease (Kanehisa et al., 2017) – Last updated 01/26/2024.
- Disgenet (Piñero et al., 2020) – Last updated October 2021.
- MalaCards (Rappaport et al., 2017) – version 5.20, last updated on 04/26/2024.
- Million Veteran Program (MVP) AUD 2019 Publication (Kranzler et al., 2019).
- MVP AUD 2023 Publication (Zhou et al., 2023).
- Prior GWAS analysis of AI cohort (Peng et al., 2024, 2019, 2017).
- Molecular Targets of Ethanol (Abrahao et al., 2017; Edenberg et al., 2010; Harris et al., 2008; Kendler et al., 2011; Koyama et al., 2007; Moore et al., 1990; Sanchez-Roige et al., 2019; Schuckit et al., 2005; You et al., 2019).

Notably some genes overlapped between sources, in this case all duplicates were removed when combining genes from each source.

Method of Attainment and Genes Attained:

- Diseases 2.0 Database: Looked up Alcohol Use Disorder Entry.

ADH1B, ADH1C, ADH4, ALDH2, CHRM2, COMT, DRD2, DRD3, GABRA2, GABRG3, HTR2A, OPRM1, SLC6A3, SLC6A4, TAS2R16.

- Kegg Disease: Alcohol Dependence entry.

ALDH2, ADH1B, ADH1C, GABRA2, HTR2A, TAS2R16.

- Disgenet: Alcohol Use Disorder entry (Filtered by score >= 0.3). Additionally, used Alcohol Intoxication, Chronic entry (Filtered by score >= 0.3).

AUD: ADH1B, ADH1C, ADH4, ALDH2, BAG3, CCKAR, CDK20, CHRNA3, CHRNA5, COMT, CYP2E1, ERCC1, FSHB, GABBR1, GABRA2, GABRG2, GATA4, GGT1, GHSR, GPHN, HTR2A, HTR3A, HTR4, LHB, MCM5, MIR146A, NPY, NPY2R, OPRM1, PDE4B, PDYN, PPM1G, SAT1, SHBG, SLC6A4, SNCA, ST18, TACR1, TACR3, TAS2R13, TAS2R16, TAS2R38, TBX19, TF, TPH2, VWF

AD: ABO, ACE, ADCY5, ADCY7, ADH1A, ADH1B, ADH1C, ADH4, ADH5, ADH6, ADH7, ADRA2A, AGO1, AGO2, AKR1A1, AKR1C3, AKR1C4, ALDH1A1, ALDH2, ALDH3B2, ALK, ANAPC1, ANKK1, ANKRD7, APOE, AR, ARC, ARSA, ASTN1, BABRB3, BAG3, BDNF, BHMT, C1D, CALCA, CAMK2A, CAMK4, CARS1, CARTPT, CASC4, CAT, CCK, CCKAR, CDH10, CDH11, CDH12, CDH13, CDH15, CDH18, CDH5, CDH8, CDH9, CDK20, CFTR, CHRM2, CHRNA3, CHRNA4, CHRNA5, CHRNB3, CHRNB4, CLOCK, CNR1, CNRN2, CNTN4, CNTN6, CNTNAP2, COMT, CREB1, CRH, CRHBP, CRHR1, CTNNA2, CXCL8, CYP2A13, CYP2B6, CYP2E1, CYTL1, DBH, DKK2, DPYSL2, DRD1, DRD2, DRD3, DRD4, DSCAML1, DTNBP1, DUSP8, ECHS1, EGF, EGFR, EP300, EPHA8, EPHX1, FABP2, FGAP, FHSB, FKBP5, FOLR1, FTO, FYN, GABBR1, GABRA1, GABRA2, GABRA5, GABRA6, GABRB1, GABRB2, GABRG1, GABRG2, GABRG3, GABRR1, GABRR2, GAD1, GAD2, GAL, GALR1, GALR2, GALR3, GAP43, GAPDH, GATA4, GEMIN4, GGH, GGT1, GH1, GHS, GHSR, GLI2, GLUL, GNB3, GPHN, GRIK1, GRIK3, GRIN1, GRIN2A, GRIN2B, GRM1, GRM2, GRM3, GRM7, GRM8, GSTM1, HAMP, HDAC2, HERPUD1, HLA-DRA, HMGB1, HNMT, HOMER1, HTR1A, HTR1B, HTR2A, HTR2C, HTR3A, HTR3B, HTR7, IL10, IL17A, IL1A, IL1B, IL1R1, IL1RN, IL6, IPO11, KANK1, KCNJ6, KIAA0040, KLF11, KPNA3, LEP, LHB, LILRA1, LINC02694, LRP8, MAOA, MAOB, MBP, MGLL, MIR382, MMP2, MMP9, MOBP, MOG, MPDZ, MTHFR, NAP1L4, NAT1, NCAM1, NEUROD2, NFKB1, NKAIN1, NKAIN2, NLGN4X, NMUR2, NPS, NPSR1, NPY, NPY2R, NPY5R, NQO2, NR4A2, NRDC, NRXN3, NTRK2, NTS, NTSR1, OPRD1, OPRK1, OPRL1, OPRM1, OSBPL5, OXT, PCDH10, PCDH12, PDE10A, PDE4B, PDYN, PENK, PER3, PHF3, PHLDA2, PIK3R1, PKNOX2, POMC, PPP1R1B, PTK2B, PTP4A1, RACK1, RASGRF2, REN, RFC1, RFX4, RGS4, SAT1, SDHAF3, SEMA5A, SERINC2, SGCE, SGIP1, SHBG, SIGMAR1, SLC17A5, SLC18A2, SLC1A2, SLC22A18, SLC29A1, SLC46A1, SLC6A1, SLC6A2, SLC6A3, SLC6A4, SLC6A5, SLC6A9, SLCO3A1, SNCA, SNORA54, SNRNP70, SPG21, SRD5A1, SRD5A2, ST18, STON2, TAC1, TACR1, TACR3, TAGLN3, TAS2R16, TAS2R38, TBX19, TESK2, TF, TFAP2B, TH, THEMIS, THSD7B, TIPARP, TKT, TP53, TPH1, TRH, TTC12, UBAP2, VWF, XRCC5, ZCCHC14, ZNF699

- MalaCards: looked up AUD entry and AD entry.

AUD Entry:

ABCB7, ACE, ADH1B, ADH1C, ADH4, ADH7, AFP, AKR1A1, ALB, ALDH2, ALDH9A1, ANKK1, ATP12A, ATP4A, AVP, BDNF, CBS, CD4, COMT, CREB1, CRH, CRHBP, CRHR1, CRP, CYP2E1, DRD2, DRD3, DRD4, F2, GABRA2, GAL, GGT1, GPT, GPT2, GRIN2B, HFE, IL1B, IL6, INS, MAOA, MIR106B, MIR140, MIR181B1, MIR196A1, MIR199A1, MIR21, MIR214, MIR223, MIR412, MIR486-1, MIR9-1, NPY, OGN, OPRM1, OXT, PDYN, POMC, SLC17A5, SLC6A3, SLC6A4, TF, TLR4, TNF

AD Entry:

ACE, ADCY7, ADH1A, ADH1B, ADH1C, ADH4, ADH5, ADH6, ADH7, AKR1A1, ALDH1A1, ALDH1B1, ALDH2, ALDH9A1, ANKK1, AUTS2, BDNF, CCK, CCKAR, CCKBR, CDH13, CHRM2, CHRNA2, CHRNA3, CHRNA4, CHRNA5, CHRNA6, CHRNB2, CHRNB3, CHRNB4, CNR1, COMT, CREB1, CRH, CRHR1, CYP2E1, DBH, DRD1, DRD2, DRD3, DRD4, DRD5, ESR1, FAAH, FTO, FYN, GABBR1, GABRA1, GABRA2, GABRA4, GABRA5, GABRA6, GABRB1, GABRB2, GABRB3, GABRG1, GABRG2, GABRG3, GABRR1, GABRR2, GAD1, GAD2, GATA4, GGT1, GHRL, GHSR, GPT, GRIK1, GRIK3, GRIN1, GRIN2A, GRIN2B, GRM2, GRM5, GRM8, HCRT, HOMER1, HOMER2, HTR1A, HTR1B, HTR2A, HTR2C, HTR3A, HTR3B, HTR7, IL1A, IL1B, IL1RN, JUN, KCNJ6, KIAA0040, LEP, LOC126807122, MAOA, MAOB, MMP9, MPDZ, MTHFR, NFKB1, NGF, NPY, NPY1R, NPY2R, NPY5R, NR4A2, NRXN3, NTRK2, OPRD1, OPRK1, OPRM1, OXT, OXTR, PDYN, PECR, PHF3, PKNOX2, PNOC, PNPLA3, POMC, PPARA, PPARG, PRL, RCBTB1, SDHAF3, SERINC2, SGCE, SGIP1, SLC17A5, SLC18A2, SLC19A3, SLC1A2, SLC6A2, SLC6A3, SLC6A4, SNCA, TACR1, TAS2R16, TGFB1, TH, TNF, TP53, TPH1, TPH2, TSPO, TTC12, ZNF699

- MVP AUD 2019 Publication: used genes listed in Figure 1.

ADH1B, ADH1C, ADH4, ADH6, ALDH2, BAHCC1, BRAP, DCLK2, DRD2, FTO, GCKR, IGF2BP1, ISL1, KCTD16, KLB, MICB, PKHD1, PPP1R3B, RBX1, SCN8A, SIX3, SLC39A8, TSPAN5, VRK2

- MVP AUD 2023 Publication: Supplemental Table 1. Nearest Gene column in excel sheet (column N). Recorded every gene that was intronic, exonic, or UTR3. Attained a total of 56 genes.

ACSS3, ADGRL3, ADH1B, ALMS1, ARID4A, BRAP, BRD3, CACNA1C, CACNA1E, CADM1, CADM2, CALN1, CELF2, CNOT4, CSMD1, CSMD3, DAG1, DPYD, DRD2, ERI3, FTO, FUT2, GABRA4, GCKR, GINS2, HLA-C, HS6ST3, INPP4B, MAPT, MLXIPL, MYO15A, NCAM1, NF1, OPRM1, PDE4B, POR, PPP1R13B, RABGAP1L, RASIP1, RPS6KA4, RSRC1, RUNX1T1, SLC25A37, SLC39A8, SLC4A8, SLC9A8, TBX6, TCF4, THSD7B, TNRC6A, TSNARE1, VRK2, XPO7, ZIC4, ZNF536, ZNF804A.

- Prior analyses of AI cohort

ADH1A, ADH1B, ADH1C, ADH4, ADH5, ADH6, ADH7, ALDH2, ASIC2, EBI3, FSTL5, KCNK2, LINC02347, OSBPL9, PDE4C, PRKG2, ROBO2, TIA1

- Molecular Targets of Ethanol:

ADCY1, ADCY2, ADCY3, ADCY4, ADCY5, ADCY6, ADCY7, ADCY8, ADCY9, ADCY10, ADH1A, ADH1B, ADH1C, ADH4, ADH5, ADH6, ADH7, CHRNA1, CHRNA2, CHRNA3, CHRNA4, CHRNA5, CHRNA6, CHRNA7, CHRNA9, CHRNA10, CHRNB1, CHRNB2, CHRNB3, CHRNB4, CHRND, CHRNE, CHRNG, CRH, CRHR1, CRHR2, CRHBP, GABRA1, GABRA2, GABRA3, GABRA4, GABRA5, GABRA6, GABRB1, GABRB2, GABRB3, GABRD, GABRE, GABRG1, GABRG2, GABRG3, GABRP, GABRQ, GABRR1, GABRR2, GABRR3, GLRA1, GLRA2, GLRA3, GLRB, GRIA1, GRIA2, GRIA3, GRIA4, GRID1, GRID2, GRIK1, GRIK2, GRIK3, GRIK4, GRIK5, GRIN1, GRIN2A, GRIN2B, GRIN2C, GRIN2D, GRIN3A, GRIN3B, GRM1, GRM2, GRM3, GRM4, GRM5, GRM6, GRM7, GRM8, HCN1, HCN2, HCN3, HCN4, HTR3A, HTR3B, HTR3C, HTR3D, HTR3E, KCNN1, KCNN2, KCNN3, KCNN4, KCNJ3, KCNJ5, KCNJ6, KCNJ9, KCNQ1, KCNQ2, KCNQ3, KCNQ4, KCNQ5, KCNMA1, KCNMB1, KCNMB2, KCNMB3, KCNMB4, LRRC25, LRRC38, LRRC52, LRRC56, PRKCA, PRKCB, PRKCD, PRKCE, PRKCG, PRKCH, PRKCI, PRKCQ, PRKCZ, UCN1, UCN2, UCN3

##### S5. REMMA Analysis

REMMA can utilize five types of SNP matrices: additive (A), dominant (D), additive by additive (AxA), additive by dominant (AxD), and dominant by dominant (DxD) (Wang et al., 2020). Additionally, REMMA has three core epistasis functions: additive by additive (remma_epiAA), additive by dominant (remma_epiAD), and dominant by dominant (remma_epiDD). We combined these to create five configurations (Supplemental Table 7). In all cases we used the approximate test, instead of exact test due to runtime constraints. Code used to generate REMMA scripts can be found in our GitHub repository (https://github.com/staslist/Biclustering_Epistasis). The configurations were generated based on gmat project description posted on pypi (<https://pypi.org/project/gmat/2020.4.9/>). Gmat is alternative name used by author for REMMA.

Supplemental Table 7: SNP matrices column lists all the SNP matrices used during the variance computation within the wemai_multi_gmat function.

| SNP Matrices | Epistasis Test | Configuration Name |
| --- | --- | --- |
| A, AxA | remma_epiAA | aa1 |
| A, D, AxA | remma_epiAA | aa2 |
| A, D, AxA, AxD, DxD | remma_epiAA | aa3 |
| A, D, AxA, AxD, DxD | remma_epiAD | ad |
| A, D, AxA, AxD, DxD | remma_epiDD | dd |

We conducted the epistasis analysis of AI cohort using all five configurations. The results were then merged. For any SNP pair that appeared in multiple configurations the most significant p-value was selected.

##### S6. Bi-clustering of epistasis results

All the functions used to carry our bi-clustering are located in BiClustering.py. The key functions are initialize_matrices2, comp_alt, compute_k_m_parallel, compute_interval_pval_parallel, and trim_intervals. The comments in each functions provide an in-depth description of the code.

Bi-Clustering Algorithm:

1) Build the interaction matrix. Fill it with 1s (interactions) and 0s (non-interactions).
2) Evaluate the total number of tested interactions (N) and the total number of interacting pairs (n).
3) Exhaustively scan the interaction matrix using a submatrix A_i,j,a,b_ to identify interacting interval pairs enriched for SNP-SNP interactions:
   a. i,j are the starting row and column indices. Since the algorithm is exhaustive, the i,j traverse the entire interaction matrix.
   b. a and b are the dimensions of the submatrix. The maximum size of the submatrix used in this analysis was 60 x 60. Minimum size was 1 x 1. For every index (i,j) a total of 3600 submatrices were evaluated (ex: A_1,1,1,1_, A_1,1,1,2_, A_1,1,1,3_, … , A_1,1,1,60_, … , A_1,1,60,60_).
   c. For each instance of submatrix, evaluate the k=number of interactions in submatrix and m=number of tested interactions within the submatrix (the size of submatrix A)
   d. Calculate the p-value for every interval pair (submatrix) using hypergeometric distribution with four parameters: m, k, N, n
4) Select most significant interval pair, remove all (less significant) interval pairs that overlap with it, repeat the process. The remaining interval pairs will be disjoint.

##### S7. Analysis of interacting genes and/or regulatory elements

The goal of this analysis was to see whether there was any prior evidence of possible interaction for gene and/or regulatory element pairs that were determined as significantly interacting by the bi-clustering algorithm.

As mentioned in methods, when expanding core gene set, the following setting was used: the overall interaction score had to be > 0.9 and both experimental and database evidence had to be above 0.01 in StringDB (Szklarczyk et al., 2023). For GeneHancer the score threshold was 25 (Fishilevich et al., 2017).

When examining for evidence of possible interaction in StringDB post bi-clustering we used ‘medium confidence’ score threshold of > 0.4. Regarding GeneHancer when examining for evidence of regulatory element interaction score threshold of 25 was used and non-protein coding genes were ignored. Analyzing regulatory elements: i) regulatory element - gene interaction: check whether this regulatory element is known to impact expression of this gene, ii) regulatory element - gene interaction: check whether genes regulated by this regulatory element interact with this gene. iii) regulatory element – regulatory element: check whether genes regulated by these regulatory elements are known to interact. When selecting genes being impacted by regulatory elements non-protein coding genes were ignored.

Additionally, we used MSigDB curated gene sets to examine whether any gene pairs were present within the same pathways (Liberzon et al., 2015).

Finally, an internet search was conducted for every pair of genes using Google. The search term used was in the form of: ‘GENE1 and GENE2’. Such as: DPYD and TIA1. We looked for any publications that mentioned both genes interacting.

##### S8. Enrichment analysis of interacting genes.

All interacting genes were analyzed against background of expanded core gene set in Enrichr and FUMA GWAS (Chen et al., 2013; Watanabe et al., 2017).

The following libraries were used in the analysis:

Enrichr: Reactome 2024, BioPlanet 2019, WikiPathway 2024, KEGG 2021, GO Biological Process 2023, GO Cellular Component 2023, GO Molecular Function 2023

FUMA GWAS: Curated Gene Sets, All Canonical Pathways, BioCarta, KEGG, Reactome, Cell Types, GO BP, GO CC, GO MF, Wikipathways, GWAS catalog reported genes

##### S9. Enrichment analysis of genes regulated by the interacting regulatory elements.

The regulated genes were analyzed against background of all human genes in Enrichr and FUMA GWAS.

The following libraries were used in the analysis:

Enrichr: Reactome 2023, BioPlanet 2019, WikiPathway 2023, KEGG 2021, GO Biological Process 2023, GO Cellular Component 2023, GO Molecular Function 2023

FUMA GWAS: Curated Gene Sets, All Canonical Pathways, BioCarta, KEGG, Reactome, Cell Types, GO BP, GO CC, GO MF, Wikipathways, GWAS catalog reported genes

#### References

Abrahao, K.P., Salinas, A.G., Lovinger, D.M., 2017. Alcohol and the Brain: Neuronal Molecular Targets, Synapses, and Circuits. Neuron 96, 1223–1238. https://doi.org/10.1016/j.neuron.2017.10.032

Bizon, C., Spiegel, M., Chasse, S.A., Gizer, I.R., Li, Y., Malc, E.P., Mieczkowski, P.A., Sailsbery, J.K., Wang, X., Ehlers, C.L., Wilhelmsen, K.C., 2014. Variant calling in low-coverage whole genome sequencing of a Native American population sample. BMC Genomics 15, 85. https://doi.org/10.1186/1471-2164-15-85

Bucholz, K.K., Cadoret, R., Cloninger, C.R., Dinwiddie, S.H., Hesselbrock, V.M., Nurnberger, J.I., Reich, T., Schmidt, I., Schuckit, M.A., 1994. A new, semi-structured psychiatric interview for use in genetic linkage studies: a report on the reliability of the SSAGA. J. Stud. Alcohol 55, 149–158. https://doi.org/10.15288/jsa.1994.55.149

Chen, E.Y., Tan, C.M., Kou, Y., Duan, Q., Wang, Z., Meirelles, G.V., Clark, N.R., Ma’ayan, A., 2013. Enrichr: interactive and collaborative HTML5 gene list enrichment analysis tool. BMC Bioinformatics 14, 128. https://doi.org/10.1186/1471-2105-14-128

DePristo, M.A., Banks, E., Poplin, R., Garimella, K.V., Maguire, J.R., Hartl, C., Philippakis, A.A., del Angel, G., Rivas, M.A., Hanna, M., McKenna, A., Fennell, T.J., Kernytsky, A.M., Sivachenko, A.Y., Cibulskis, K., Gabriel, S.B., Altshuler, D., Daly, M.J., 2011. A framework for variation discovery and genotyping using next-generation DNA sequencing data. Nat. Genet. 43, 491–498. https://doi.org/10.1038/ng.806

Edenberg, H.J., Koller, D.L., Xuei, X., Wetherill, L., McClintick, J.N., Almasy, L., Bierut, L.J., Bucholz, K.K., Goate, A., Aliev, F., Dick, D., Hesselbrock, V., Hinrichs, A., Kramer, J., Kuperman, S., Nurnberger Jr, J.I., Rice, J.P., Schuckit, M.A., Taylor, R., Todd Webb, B., Tischfield, J.A., Porjesz, B., Foroud, T., 2010. Genome-Wide Association Study of Alcohol Dependence Implicates a Region on Chromosome 11. Alcohol. Clin. Exp. Res. 34, 840–852. https://doi.org/10.1111/j.1530-0277.2010.01156.x

Ehlers, C.L., Stouffer, G.M., Corey, L., Gilder, D.A., 2015. The clinical course of DSM-5 alcohol use disorders in young adult native and Mexican Americans. Am. J. Addict. 24, 713–721. https://doi.org/10.1111/ajad.12290

Ehlers, C.L., Wall, T.L., Betancourt, M., Gilder, D.A., 2004. The Clinical Course of Alcoholism in 243 Mission Indians. Am. J. Psychiatry 161, 1204–1210. https://doi.org/10.1176/appi.ajp.161.7.1204

Fishilevich, S., Nudel, R., Rappaport, N., Hadar, R., Plaschkes, I., Iny Stein, T., Rosen, N., Kohn, A., Twik, M., Safran, M., Lancet, D., Cohen, D., 2017. GeneHancer: genome-wide integration of enhancers and target genes in GeneCards. Database 2017, bax028. https://doi.org/10.1093/database/bax028

Gilder, D.A., Wall, T.L., Ehlers, C.L., 2004. Comorbidity of Select Anxiety and Affective Disorders With Alcohol Dependence in Southwest California Indians. Alcohol. Clin. Exp. Res. 28, 1805–1813. https://doi.org/10.1097/01.ALC.0000148116.27875.B0

Grissa, D., Junge, A., Oprea, T.I., Jensen, L.J., 2022. Diseases 2.0: a weekly updated database of disease–gene associations from text mining and data integration. Database 2022, baac019. https://doi.org/10.1093/database/baac019

Harris, R.A., Trudell, J.R., Mihic, S.J., 2008. Ethanol’s Molecular Targets. Sci. Signal. 1, re7–re7. https://doi.org/10.1126/scisignal.128re7

Heckathorn, D.D., 1997. Respondent-Driven Sampling: A New Approach to the Study of Hidden Populations*. Soc. Probl. 44, 174–199. https://doi.org/10.2307/3096941

Hesselbrock, M., Easton, C., Bucholz, K.K., Schuckit, M., Hesselbrock, V., 1999. A validity study of the SSAGA-a comparison with the SCAN. Addiction 94, 1361–1370. https://doi.org/10.1046/j.1360-0443.1999.94913618.x

Hesselbrock, V.M., Hesselbrock, M.N., Segal, B., 2003. Alcohol Dependence Among Alaskan Natives and Their Health Care Utilization. Alcohol. Clin. Exp. Res. 27, 1353–1355. https://doi.org/10.1097/01.ALC.0000080167.17411.21

Hesselbrock, V.M., Segal, B., Hesselbrock, M.N., 2000. Alcohol dependence among Alaska Natives entering alcoholism treatment: a gender comparison. J. Stud. Alcohol 61, 150–156. https://doi.org/10.15288/jsa.2000.61.150

Kalton, G., Anderson, D.W., 1986. Sampling Rare Populations. J. R. Stat. Soc. Ser. Gen. 149, 65–82. https://doi.org/10.2307/2981886

Kanehisa, M., Furumichi, M., Tanabe, M., Sato, Y., Morishima, K., 2017. KEGG: new perspectives on genomes, pathways, diseases and drugs. Nucleic Acids Res. 45, D353–D361. https://doi.org/10.1093/nar/gkw1092

Kendler, K.S., Kalsi, G., Holmans, P.A., Sanders, A.R., Aggen, S.H., Dick, D.M., Aliev, F., Shi, J., Levinson, D.F., Gejman, P.V., 2011. Genomewide Association Analysis of Symptoms of Alcohol Dependence in the Molecular Genetics of Schizophrenia (MGS2) Control Sample. Alcohol. Clin. Exp. Res. 35, 963–975. https://doi.org/10.1111/j.1530-0277.2010.01427.x

Koyama, S., Brodie, M.S., Appel, S.B., 2007. Ethanol Inhibition of M-Current and Ethanol-Induced Direct Excitation of Ventral Tegmental Area Dopamine Neurons. J. Neurophysiol. 97, 1977–1985. https://doi.org/10.1152/jn.00270.2006

Kranzler, H.R., Zhou, H., Kember, R.L., Vickers Smith, R., Justice, A.C., Damrauer, S., Tsao, P.S., Klarin, D., Baras, A., Reid, J., Overton, J., Rader, D.J., Cheng, Z., Tate, J.P., Becker, W.C., Concato, J., Xu, K., Polimanti, R., Zhao, H., Gelernter, J., 2019. Genome-wide association study of alcohol consumption and use disorder in 274,424 individuals from multiple populations. Nat. Commun. 10, 1499. https://doi.org/10.1038/s41467-019-09480-8

Li, Y., Sidore, C., Kang, H.M., Boehnke, M., Abecasis, G.R., 2011. Low-coverage sequencing: Implications for design of complex trait association studies. Genome Res. 21, 940–951. https://doi.org/10.1101/gr.117259.110

Liberzon, A., Birger, C., Thorvaldsdóttir, H., Ghandi, M., Mesirov, J.P., Tamayo, P., 2015. The Molecular Signatures Database (MSigDB) hallmark gene set collection. Cell Syst. 1, 417–425. https://doi.org/10.1016/j.cels.2015.12.004

Moore, S.D., Madamba, S.G., Siggins, G.R., 1990. Ethanol diminishes a voltage-dependent K+ current, the M-current, in CA1 hippocampal pyramidal neurons in vitro. Brain Res. 516, 222–228. https://doi.org/10.1016/0006-8993(90)90922-X

Muhib, F.B., Lin, L.S., Stueve, A., Miller, R.L., Ford, W.L., Johnson, W.D., Smith, P.J., 2001. A Venue-Based Method for Sampling Hard-to-Reach Populations. Public Health Reports® 116, 216–222. https://doi.org/10.1093/phr/116.S1.216

Peng, Q., Bizon, C., Gizer, I.R., Wilhelmsen, K.C., Ehlers, C.L., 2019. Genetic loci for alcohol-related life events and substance-induced affective symptoms: indexing the “dark side” of addiction. Transl. Psychiatry 9, 1–12. https://doi.org/10.1038/s41398-019-0397-6

Peng, Q., Gilder, D.A., Bernert, R.A., Karriker-Jaffe, K.J., Ehlers, C.L., 2024. Genetic factors associated with suicidal behaviors and alcohol use disorders in an American Indian population. Mol. Psychiatry 29, 902–913. https://doi.org/10.1038/s41380-023-02379-3

Peng, Q., Gizer, I.R., Wilhelmsen, K.C., Ehlers, C.L., 2017. Associations Between Genomic Variants in Alcohol Dehydrogenase Genes and Alcohol Symptomatology in American Indians and European Americans: Distinctions and Convergence. Alcohol. Clin. Exp. Res. 41, 1695–1704. https://doi.org/10.1111/acer.13480

Piñero, J., Ramírez-Anguita, J.M., Saüch-Pitarch, J., Ronzano, F., Centeno, E., Sanz, F., Furlong, L.I., 2020. The DisGeNET knowledge platform for disease genomics: 2019 update. Nucleic Acids Res. 48, D845–D855. https://doi.org/10.1093/nar/gkz1021

Rappaport, N., Fishilevich, S., Nudel, R., Twik, M., Belinky, F., Plaschkes, I., Stein, T.I., Cohen, D., Oz-Levi, D., Safran, M., Lancet, D., 2017. Rational confederation of genes and diseases: NGS interpretation via GeneCards, MalaCards and VarElect. Biomed. Eng. OnLine 16, 72. https://doi.org/10.1186/s12938-017-0359-2

Sanchez-Roige, S., Fontanillas, P., Elson, S.L., Team, T. 23andMe R., Gray, J.C., de Wit, H., Davis, L.K., MacKillop, J., Palmer, A.A., 2019. Genome-wide association study of alcohol use disorder identification test (AUDIT) scores in 20 328 research participants of European ancestry. Addict. Biol. 24, 121–131. https://doi.org/10.1111/adb.12574

Schuckit, M.A., Smith, T.L., Anthenelli, R., Irwin, M., 1993. Clinical course of alcoholism in 636 male inpatients. Am. J. Psychiatry 150, 786–792. https://doi.org/10.1176/ajp.150.5.786

Schuckit, M.A., Wilhelmsen, K., Smith, T.L., Feiler, H.S., Lind, P., Lange, L.A., Kalmijn, J., 2005. Autosomal Linkage Analysis for the Level of Response to Alcohol. Alcohol. Clin. Exp. Res. 29, 1976–1982. https://doi.org/10.1097/01.alc.0000187598.82921.27

Szklarczyk, D., Kirsch, R., Koutrouli, M., Nastou, K., Mehryary, F., Hachilif, R., Gable, A.L., Fang, T., Doncheva, N.T., Pyysalo, S., Bork, P., Jensen, L.J., von Mering, C., 2023. The STRING database in 2023: protein–protein association networks and functional enrichment analyses for any sequenced genome of interest. Nucleic Acids Res. 51, D638–D646. https://doi.org/10.1093/nar/gkac1000

Van der Auwera, G.A., Carneiro, M.O., Hartl, C., Poplin, R., del Angel, G., Levy-Moonshine, A., Jordan, T., Shakir, K., Roazen, D., Thibault, J., Banks, E., Garimella, K.V., Altshuler, D., Gabriel, S., DePristo, M.A., 2013. From FastQ Data to High-Confidence Variant Calls: The Genome Analysis Toolkit Best Practices Pipeline. Curr. Protoc. Bioinforma. 43, 11.10.1-11.10.33. https://doi.org/10.1002/0471250953.bi1110s43

Wang, D., Tang, H., Liu, J.-F., Xu, S., Zhang, Q., Ning, C., 2020. Rapid epistatic mixed-model association studies by controlling multiple polygenic effects. Bioinformatics 36, 4833–4837. https://doi.org/10.1093/bioinformatics/btaa610

Watanabe, K., Taskesen, E., van Bochoven, A., Posthuma, D., 2017. Functional mapping and annotation of genetic associations with FUMA. Nat. Commun. 8, 1826. https://doi.org/10.1038/s41467-017-01261-5

You, C., Savarese, A., Vandegrift, B.J., He, D., Pandey, S.C., Lasek, A.W., Brodie, M.S., 2019. Ethanol acts on KCNK13 potassium channels in the ventral tegmental area to increase firing rate and modulate binge–like drinking. Neuropharmacology 144, 29–36. https://doi.org/10.1016/j.neuropharm.2018.10.008

Zhou, H., Kember, R.L., Deak, J.D., Xu, H., Toikumo, S., Yuan, K., Lind, P.A., Farajzadeh, L., Wang, L., Hatoum, A.S., Johnson, J., Lee, H., Mallard, T.T., Xu, J., Johnston, K.J.A., Johnson, E.C., Nielsen, T.T., Galimberti, M., Dao, C., Levey, D.F., Overstreet, C., Byrne, E.M., Gillespie, N.A., Gordon, S., Hickie, I.B., Whitfield, J.B., Xu, K., Zhao, H., Huckins, L.M., Davis, L.K., Sanchez-Roige, S., Madden, P.A.F., Heath, A.C., Medland, S.E., Martin, N.G., Ge, T., Smoller, J.W., Hougaard, D.M., Børglum, A.D., Demontis, D., Krystal, J.H., Gaziano, J.M., Edenberg, H.J., Agrawal, A., Zhao, H., Justice, A.C., Stein, M.B., Kranzler, H.R., Gelernter, J., 2023. Multi-ancestry study of the genetics of problematic alcohol use in over 1 million individuals. Nat. Med. 29, 3184–3192. https://doi.org/10.1038/s41591-023-02653-5
